# Hospital AI and Robotics Adoption, Access Inequality, and County Mortality: A National Study Across 3,143 U.S. Counties

**DOI:** 10.64898/2026.03.10.26347904

**Authors:** Aaron Johnson, David Gefen, Teresa Harrison

## Abstract

Hospital artificial intelligence (AI) and robotics are spreading unevenly across the United States, yet national evidence on how these technologies are associated with hospital performance and population health remains limited. This observational study linked the 2024 American Hospital Association Annual Survey, capturing calendar-year 2023 adoption across 6,166 hospitals, to CMS, CDC, and County Health Rankings outcomes for 3,143 U.S. counties. Conditioning on 2019 baseline performance, hospitals using AI for staff scheduling had roughly 4% higher adherence to the SEP-1 sepsis bundle, and hospitals using AI for routine-task automation had approximately 5.1% lower 30-day pneumonia mortality. At the county level, the clearest timing-aligned outcome was contemporaneous hospital-setting mortality (2023): any access to workflow AI hospitals was associated with 25.5 fewer hospital deaths per 100,000 residents under cross-fitted doubly robust estimation (9.9% lower; non-cross-fitted estimate: 16.9 fewer per 100,000). A historical county benchmark (County Health Rankings premature mortality, 2020–2022) showed a concordant pattern, with the cross-fitted estimate corresponding to 975.8 fewer years of potential life lost per 100,000 before age 75 (*p* < 0.001), though this outcome predates the 2023 adoption measurement and should be interpreted as a lagged benchmark rather than a contemporaneous association. Workflow AI access was also associated with about 206 fewer preventable hospital stays per 100,000 residents (7.2% lower). Robotics showed narrower and more mixed associations. Yet access reflected a substantial digital divide: only 65.8% of Americans lived within 30 minutes of AI-enabled care, leaving about 114.6 million people outside that catchment (AI access Gini = 0.740), and despite a 56% increase in AI-enabled hospitals between 2022 and 2024, distributional inequality persisted. Overall, the evidence is consistent with adjusted associations between workflow AI access and better rescue outcomes where adoption occurs.

**Significance Statement:** In this study of 6,166 hospitals and 3,143 U.S. counties, hospitals using AI for staff scheduling had roughly 4% higher adherence to the recommended sepsis care bundle, and cross-fitted doubly robust estimation associated workflow AI with 25.5 fewer contemporaneous hospital deaths per 100,000 residents (9.9% lower than comparable unexposed counties; non-cross-fitted baseline: 16.9 fewer per 100,000). A concordant historical benchmark (premature mortality, 2020–2022) and a 7.2% reduction in preventable hospital stays reinforce the pattern. Yet about 114.6 million Americans live beyond a 30-minute drive of any hospital using AI. Only 65.8% of Americans lived within 30 minutes of AI-enabled care (AI access Gini = 0.740), and despite a 56% increase in AI-enabled hospitals between 2022 and 2024, distributional inequality did not improve. The contribution is not a blanket claim that AI saves lives; it is a national estimate of health disparities created when hospital technologies diffuse unevenly, suggesting that extending AI-enabled care to underserved areas may represent a high-return intervention.

## 1 Introduction

Hospital artificial intelligence (AI) and robotics are being deployed unevenly across the United States. Large, digitally mature health systems tend to adopt these tools first, while many rural communities remain far from AI-enabled or robotics-enabled care ^1,2^. Yet national estimates of what these technologies are worth to care delivery—and what populations lose when adoption lags—remain limited.

This gap matters because the practical promise of hospital AI and robotics is operational, not merely symbolic. Hospitals face persistent staffing strain, burnout, and coordination failures in time-sensitive care ^3,4^. While visionaries have described a more efficient and humane future driven by AI ^5,6^, empirical evidence linking specific hospital AI and robotics capabilities to rescue-process adherence, mortality, and access remains scarce. Recent work describes the uptake of generative AI in electronic health records ^7^, but national evidence on measured healthcare value and uneven access is still emerging. This intervention-oriented framing aligns with recent work arguing that algorithmic systems in social settings should be evaluated by how they change organizational decisions and downstream outcomes rather than by predictive performance alone ^8^.

This gap also has an equity dimension. Prior work has shown that algorithmic systems in health care can reproduce or amplify disparities when design choices, implementation resources, or access are uneven ^9,10^. The present question is therefore not only whether hospitals that adopt AI perform differently, but whether uneven diffusion creates another layer of geographic inequality in who can plausibly reach AI-enabled care.

To the best of our knowledge, this is the first national analysis to estimate both the healthcare value and the access inequality of hospital AI and robotics while conditioning on historical baseline performance. The study links the 2024 American Hospital Association (AHA) Annual Survey, covering 6,166 hospitals, to Centers for Medicare & Medicaid Services (CMS), County Health Rankings, and CDC mortality outcomes for 3,143 U.S. counties. It distinguishes between cognitive workflow AI tools that automate information processing and robotics that support physical procedures. Here, “wealth-proxy bias” means that technology adoption can stand in for pre-existing hospital resources, market power, or quality rather than for technology value itself. Conditioning on 2019 baseline outcomes therefore asks whether 2023 adopters outperform otherwise similar non-adopters relative to their own historical starting points, rather than merely reflecting the fact that better-resourced hospitals adopt first.

At the county level, the empirical median of workflow AI exposure is zero, so the main estimand is most cautiously read as an extensive-margin contrast between any access and none, supplemented by continuous-treatment and spatial robustness checks. The primary estimates reported in the body of this paper therefore emphasize interpretable adjusted mean differences on the observed scale, while Double Machine Learning (DML), Targeted Maximum Likelihood Estimation (TMLE), overlap weighting, and spatial models are used as triangulating robustness checks rather than as replacements for that primary estimand.

### Theoretical Framework

We interpret the results through the Resource-Based View (RBV) of the firm ^11,12^. In this setting, hospital AI workflow tools and robotics can be understood as organizational resources that may improve performance when hospitals can integrate them into care delivery. RBV is used here as an interpretive theoretical lens for understanding value realization after adoption rather than as a second empirical claim about diffusion.

### Research Questions

#### RQ1 (Mechanism)

To what extent are the hospital AI workflow capabilities and robotics observed in the AHA survey associated with rescue and process outcomes (SEP-1, pneumonia mortality, and procedural safety) net of baseline performance?

#### RQ2 (Scaling)

Is access-weighted exposure to AI-enabled and robotics-enabled hospitals associated with lower county-level premature mortality?

#### RQ3 (Adoption/Equity)

How unevenly are AI and robotics adopted across counties, and what access penalty does that create for rural populations?

## 2 Results

### Sample and Adoption Landscape

Our analytical sample included 6,166 U.S. hospitals in the 2024 AHA Annual Survey and all 3,143 U.S. counties after county-level aggregation. Adoption was uneven: approximately 31% of counties had at least one hospital using AI for any purpose, while robotics adoption was present in about 45% of counties. Throughout the county analyses, the focal workflow AI exposure is AI for automating routine tasks (AHA item MO14), and the focal robotics exposure is in-hospital robotics (AHA item MO21). These measures are distinct from the hospital-level staff-scheduling AI measure used in the SEP-1 mechanism analysis. Workflow AI and procedural robotics are therefore treated as related but non-interchangeable technologies (Table 1). Fig. 1 provides a national three-panel overview of continuous access, policy-threshold access, and distributional inequality.

**Table 1.**
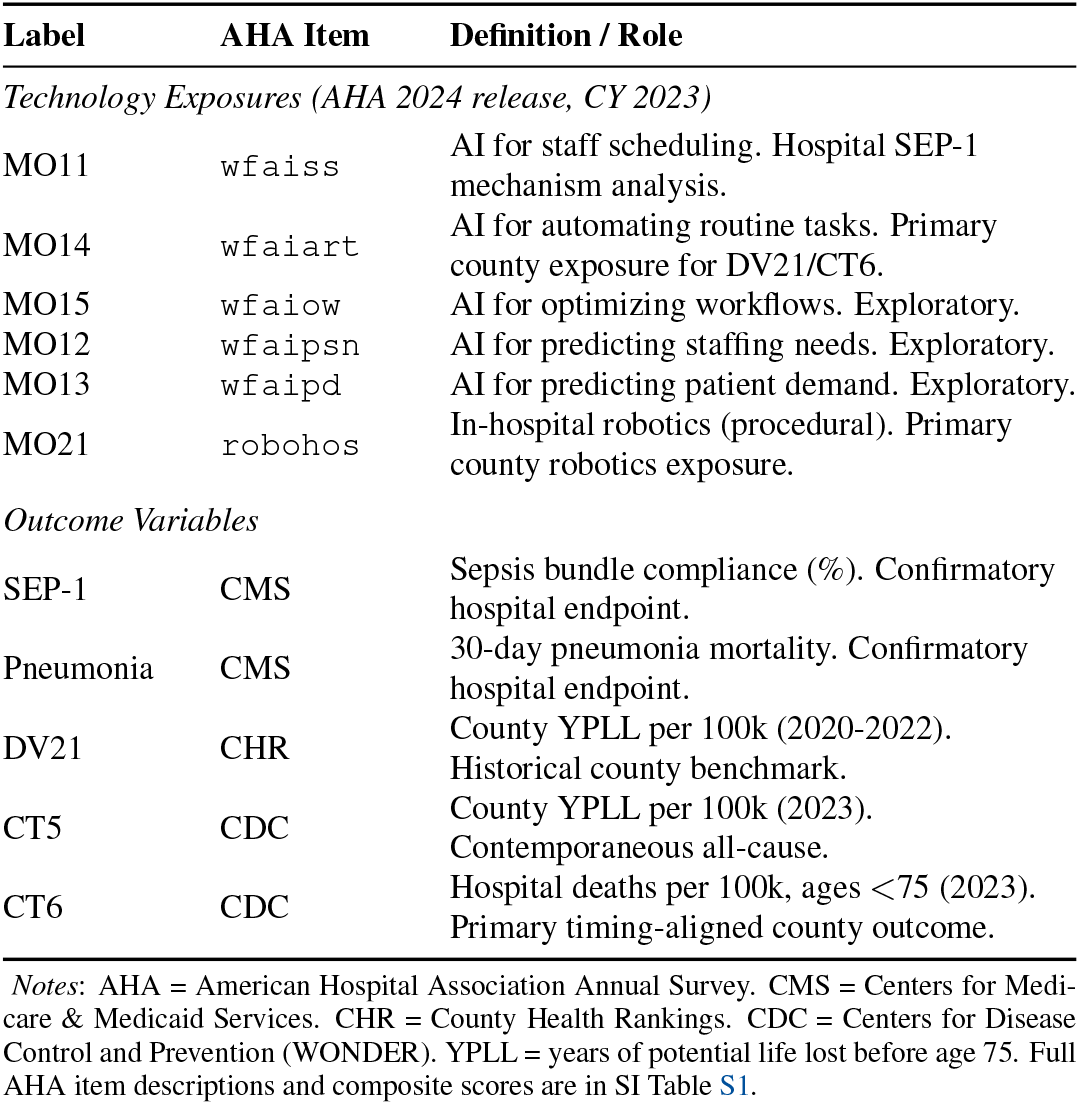
Variable Dictionary: Key AHA Technology Measures and Outcome Definitions.

**Fig. 1.**
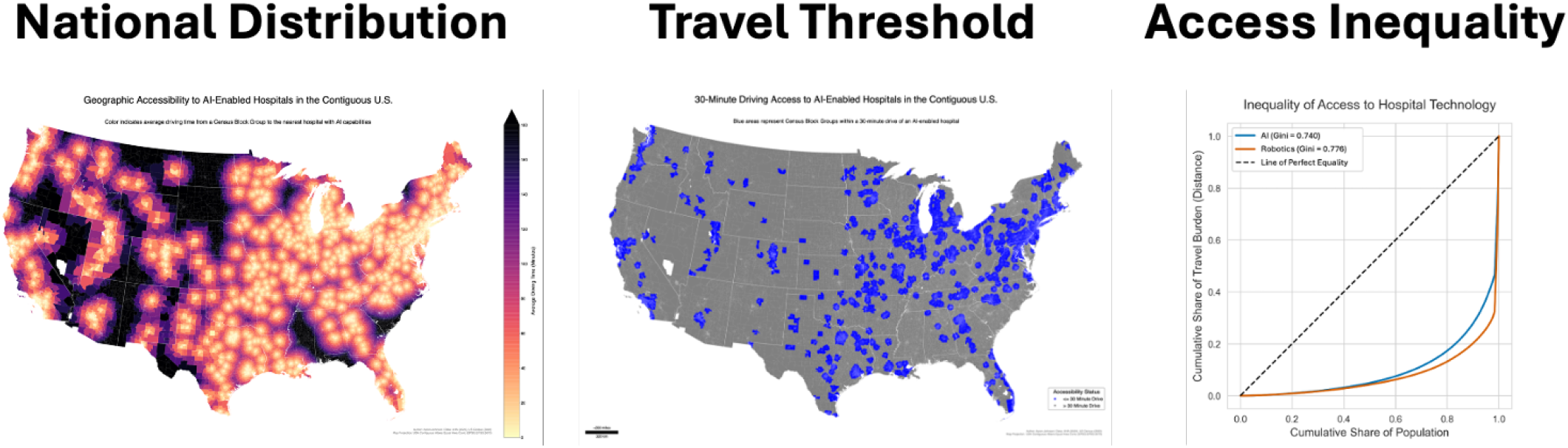
National Geographic Access and Inequality in AI- and Robotics-Enabled Hospital Access (2023). **Panel A:** Continuous access map using Census block-group proxy drive time to the nearest AI-enabled hospital in the contiguous U.S., where AI-enabled status is defined using the workflow AI routine-task automation measure (AHA item MO14, AI for automating routine tasks) aligned with the primary county mortality models. **Panel B:** Binary policy-threshold map indicating block groups within a 30-minute drive of an AI-enabled hospital; in 2023, 65.8% of the U.S. population was within 30 minutes, and approximately 114.6 million remained outside this access threshold. **Panel C:** Lorenz curves for population-weighted travel burden to AI- and robotics-enabled hospitals, showing high but differential inequality (AI Gini = 0.740; Robotics Gini = 0.776). Together, the three panels visualize national technology access deserts versus oases and their distributional consequences.

### 2.1 RQ1: Hospital-Level Mechanism (Confirmatory)

To reduce the risk that adoption simply proxies for institutional wealth or the baseline performance of already high-performing hospitals, the hospital analysis used Augmented Inverse Probability Weighting (AIPW) estimators ^13^ (see SI Section S7 for full specification and triangulation) while controlling for bed size, ownership, teaching status, county-level social determinants, and **baseline 2019 performance**. AIPW is the primary estimator because adoption is strongly selected on observable characteristics, and its doubly robust structure remains consistent if either the propensity model or the outcome model is correctly specified. Throughout Tables 2–3, the average treatment effect (ATE) is the adjusted mean difference between adopters and otherwise comparable non-adopters in the native units of each outcome; positive ATEs indicate higher values on beneficial measures, whereas negative ATEs indicate lower rates on adverse outcomes.

**Table 2.** Primary Hospital-Level AIPW Results (Contemporaneous 2023 Data)

| Treatment (CY 2023) | Outcome (CY 2023) | ATE | 95% CI |
| --- | --- | --- | --- |
| *Confirmatory endpoints (pre-specified)* |  |  |  |
| AI: Staff Scheduling | SEP-1 sepsis bundle compliance | +2.24 | [1.03, 3.44] |
| AI: Routine Tasks | Pneumonia mortality measure | -0.87 | [-1.35, -0.40] |
| *Exploratory outcomes* |  |  |  |
| AI: Patient Demand | ED wait time (minutes) | +5.30 | [2.09, 10.14] |
| Robotics | Accidental puncture (PSI-15) | +0.03<sup>a</sup> | [0.01, 0.05] |

*Note:* Estimates derive from hospital-level AIPW models ( $N = 6,166$ ) controlling for 2019 baseline performance, bed size, ownership, adjusted patient days, and state fixed effects. The ATE is the adjusted mean difference between adopters and comparable non-adopters in the native units of each outcome. Confirmatory endpoints were pre-specified based on the operational-rescue hypothesis; exploratory outcomes are reported with FDR correction in Table 3.
<sup>a</sup> PSI-15 values are inverted such that positive coefficients indicate improved safety (fewer punctures).

**Table 3.** Hospital-Level AIPW Results: Selected FDR-Corrected Significant Findings.

| Treatment | Outcome | ATE | SE | P-Value |
| --- | --- | --- | --- | --- |
| Robotics | 30-day mort. CABG | -0.24 | 0.08 | <0.01 |
| Robotics | 30-day mort. stroke | -0.30 | 0.11 | <0.01 |
| Robotics | Would recommend | 3.14 | 0.47 | <0.01 |
| Robotics | ED time (non-psych) | 8.65 | 1.68 | <0.01 |
| Robotics | MRSA SIR | 0.08 | 0.03 | 0.01 |
| AI Scheduling | 30-day mort. COPD | -0.22 | 0.09 | 0.02 |
| AI Staffing | 30-day mort. HF | -0.40 | 0.12 | <0.01 |
| AI Staffing | 30-day mort. pneumonia | -0.60 | 0.19 | <0.01 |
| AI Staffing | Pressure ulcer (PSI-03) | -0.07 | 0.03 | 0.01 |
| AI Staffing | Sepsis care (SEP-1) | 2.24 | 0.93 | 0.02 |
| AI Staffing | Flu vaccination | 5.24 | 0.85 | <0.01 |
| AI Staffing | VTE prophylaxis | 8.29 | 1.01 | <0.01 |
| AI Staffing | 30-day readm. pneumonia | -0.16 | 0.05 | <0.01 |
| AI Staffing | 30-day readm. hosp-wide | -0.11 | 0.03 | <0.01 |
| AI Patient Demand | 30-day mort. pneumonia | -0.77 | 0.14 | <0.01 |
| AI Patient Demand | Fall fracture (PSI-08) | -0.005 | 0.002 | 0.04 |
| AI Patient Demand | Flu vaccination | 6.77 | 0.76 | <0.01 |
| AI Patient Demand | 30-day readm. HF | -0.20 | 0.06 | <0.01 |
| AI Workflows | 30-day mort. pneumonia | -0.58 | 0.15 | <0.01 |
| AI Workflows | Fall fracture (PSI-08) | -0.005 | 0.002 | 0.03 |
| AI Workflows | Sepsis 3hr bundle | 1.43 | 0.61 | 0.02 |
| AI Workflows | Sepsis care (SEP-1) | 2.43 | 0.92 | <0.01 |
| AI Workflows | Flu vaccination | 6.47 | 0.81 | <0.01 |
| AI Workflows | 30-day readm. pneumonia | -0.17 | 0.05 | <0.01 |

Notes: FDR-corrected significant findings ( $q < 0.05$ ) from comprehensive hospital-level AIPW analysis ( $N = 6,166$ hospitals, 500 bootstrap iterations). ATE = average treatment effect, interpreted here as the adjusted mean difference between adopters and non-adopters in the native units of each outcome (for example, percentage points for SEP-1-type measures and minutes for ED throughput). All estimates are FDR-significant with 95% CIs excluding zero. Relative changes range from -13.9% (pressure ulcers) to +11.1% (MRSA). All models control for 2019 baseline performance, hospital characteristics, and state fixed effects. Full results with confidence intervals are available in the SI.

### Operational Rescue: Sepsis and Pneumonia

Outcomes dependent on timely intervention (“rescue”) showed the strongest robust associations (Table 2).

- **AI for Staff Scheduling:** Associated with a **2.24-point increase** in adherence to the SEP-1 sepsis bundle (*p <* 0.05).
- **AI for Routine Tasks:** Associated with a **0.87-point reduction** in the hospital-level pneumonia mortality measure (ATE ≈ − 0.87; *p <* 0.05).

These findings are consistent with the idea that AI tools may improve workforce allocation, ensuring staff are available to execute time-sensitive protocols. It is possible that, by reducing administrative friction, these tools allow clinical teams to focus cognitive resources on patient stabilization. We interpret the SEP-1 result cautiously because SEP-1 is documentation-intensive and debated as a stand-alone quality signal ^14–16^. The workflow AI pattern is not confined to SEP-1 itself: AI Workflows also showed a positive association with the sepsis 3-hour bundle component (+1.43 points; *q <* 0.05), AI Routine Tasks with lower pneumonia mortality, and MO14’s county association with CT6 was moderated by OP-18b rather than SEP-1.

### Surgical Precision (Robotics)

**Robotics adoption** was associated with a significant 0.03-point improvement in the inverted Patient Safety Indicator 15 (Accidental Puncture or Laceration) measure—that is, fewer puncture/laceration events after direction was harmonized so positive coefficients indicate better safety (*q <* 0.05). However, associations with broader vascular outcomes like Deep Vein Thrombosis (PSI-12) were not statistically significant after adjustment. This highlights the specificity of the mechanism: robotics aids procedural precision, while workflow AI aids systemic process adherence.

### Post-hoc baseline checks

To probe reverse causality (e.g., that high-performing hospitals simply adopt AI), we ran post-hoc baseline checks that regressed 2023 AI adoption (from the 2024 AHA release) on **2019 baseline outcomes**. For our primary mechanisms (AI Staffing → SEP-1; AI Routine Tasks → Pneumonia), there was **no significant association** between 2023 adoption and 2019 performance (*p >* 0.10, see SI Table S2). This supports the interpretation that the benefits observed in 2023 are not merely reflections of historical quality but are consistent with newer operational capabilities. As an additional check, we tested hospital-level outcomes with no plausible rescue-process mechanism: elective hip/knee complication rates and colonoscopy follow-up. None showed significant associations with AI or robotics adoption (SI Table S15, Panel B), supporting causal specificity.

#### Bridging hospital mechanisms to population mortality

To probe whether the proposed operational-rescue pathway extends beyond SEP-1 documentation to measurable mortality outcomes, a two-stage bridge linked the focal workflow AI measure—AI for automating routine tasks (MO14)—and robotics (MO21) to hospital mechanism metrics and then to county mortality. In the first-stage models, MO14 was associated with lower 30-day pneumonia mortality (*β* = − 0.57 per percentage-point; *q <* 0.001), and MO21 was associated with faster ED throughput (*β* = +16.2 minutes; *q <* 0.001). In the second-stage models, pneumonia mortality independently predicted county YPLL (*β* = − 59.2; *p* = 0.002). When the county outcome was contemporaneous 2023 all-cause premature mortality (CT5; YPLL per 100,000 before age 75), adding pneumonia mortality attenuated the MO14 coefficient by 20.1%, while adding the full mechanism set (SEP-1, PSI-90, pneumonia mortality, and ED throughput) attenuated MO14 by 19.8% and MO21 by 26.4%. Future bridge specifications that additionally control for hospital payer mix (Medicare/Medicaid share), safety-net designation, and nurse-to-bed staffing ratios could further test the stability of these attenuation patterns, particularly for robotics where referral-selection concerns are strongest. These bridge results complement the CT6 analysis: in the primary cross-fitted county model, workflow AI was associated with lower hospital-setting mortality (cross-fitted AIPW ATE = − 25.5 deaths per 100,000; *p <* 0.001; about 9.9% relative reduction), while a two-part model indicated that the extensive margin drives the CT6 signal (ATE = − 15.7) and the intensive margin among adopters was approximately null (OLS slope = −0.047, *p* = 0.50; GPS slope = − 0.009, *p* = 0.91). Separately, higher SEP-1 compliance was independently associated with lower CT6 (ATE = − 9.3; *p* = 0.012), and poorer ED throughput independently predicted higher CT6 (ATE = +28.3; *p* = 0.004). Because the significant OP-18b *×* MO14 interaction appears on CT6 rather than on SEP-1 itself, the mechanism-consistent pattern is not reducible to SEP-1 documentation alone.

### Contemporaneous Hospital-Setting Mortality Associations (CT6)

To test whether the proposed operational-rescue pathway is detectable in an outcome measured in the same year as technology exposure, we constructed CT6, an age-adjusted hospital death rate per 100,000 county residents (ages *<* 75) for 2023 from CDC WONDER mortality tabulated by county of residence, age band, and hospital place of death. Counties with suppressed CT6 cells after tabulation were excluded rather than imputed in the bridge analyses; of 3,143 counties, 247 (7.9%) were excluded due to CDC suppression (death counts 1–9), predominantly rural counties with populations below 20,000. Excluded counties had lower median populations (8,412 vs. 27,891) and higher baseline YPLL (9,847 vs. 8,562), suggesting that suppression-related selection, if anything, removes counties where health outcomes are worse, biasing remaining estimates toward the null. Small-area estimation or Bayesian partial-pooling approaches could recover estimates for these suppressed cells in future work; the present analysis takes the conservative approach of complete-case exclusion rather than imputation under untested distributional assumptions. In the primary county sample, the untreated mean CT6 was 257.5 hospital deaths per 100,000 county residents. Under 5-fold cross-fitted doubly robust estimation, counties with any exposure to workflow AI for routine task automation (MO14) exhibited 25.5 fewer hospital deaths per 100,000 (95% CI [ − 39.7, − 11.3]; *p <* 0.001), a 9.9% reduction relative to the control mean (effective sample size = 409.1; 99th-percentile weight = 19.6). The non-cross-fitted AIPW baseline yielded a more conservative estimate of − 16.9 per 100,000 (95% CI [ − 29.9, − 6.2]; *p* = 0.008; 6.6% reduction). Robotics exposure (MO21) was directionally similar (cross-fitted: 25.8 per 100,000; non-cross-fitted: − 13.0; 95% CI [ − 27.9, 0.1]; *p* = 0.060), while naïve OLS slopes were weakly positive for robotics and near-zero for workflow AI (Table 4), underscoring why adjustment for baseline quality and selection matters in this setting. Counties above the median SEP-1 also showed significantly lower CT6 (ATE = − 9.3; 95% CI [ − 15.4, − 2.7]; *q* = 0.032), and poorer ED throughput independently predicted higher CT6 (ATE = +28.3; 95% CI [10.7, 46.9]; *q* = 0.032). Because SEP-1 is partly documentation-sensitive and debated as a stand-alone quality measure, we do not interpret that result in isolation; instead, we read it jointly with pneumonia mortality, the sepsis 3-hour bundle, preventable hospital stays, ED throughput, and CT6^14–16^. The OP-18b *×* MO14 interaction is positive and FDR-significant (*β* = +0.0037; *p* = 0.004; *q* = 0.011). Because lower OP-18b indicates faster ED throughput, this implies that MO14’s protective association is strongest in counties with efficient baseline ED operations (low OP-18b) and weakens as throughput deteriorates (higher OP-18b). Because this moderation appears on CT6, a CDC-derived hospital-death outcome rather than on SEP-1 itself, the mechanism-consistent pattern is not reducible to SEP-1 documentation alone. Together, these results suggest that the clearest timing-aligned county signal is hospital-setting mortality, where the proposed operational pathway is most direct.

**Table 4.** Mechanism-consistent evidence linking workflow AI and robotics to contemporaneous county hospital mortality (CT6, 2023)

| Panel | Model / Test | N | Predictor | OLS $\beta$ (p) | AIPW ATE | 95% CI | Cross-fitted ATE | Cross-fitted 95% CI | q |
| --- | --- | --- | --- | --- | --- | --- | --- | --- | --- |
| A | Direct tech effects (Main) | 2,896 | MO14 | +0.04 (0.54) | −16.9 | [−29.9, −6.2] | −**25.5** | [−**39.7**, −**11.3**] | 0.032 |
|  |  | 2,896 | MO21 | +0.10 (0.05) | −13.0 | [−27.9, 0.1] | −**25.8** | [−**40.1**, −**11.5**] | 0.096 |
| B | Mechanism: SEP-1 → CT6 | 2,263 | SEP-1 | −0.26 (0.014) | −9.3 | [−15.4, −2.7] | — | — | 0.032 |
| C | Direct effect: OP-18b → CT6 | 2,263 | OP-18b | +0.18 (0.003) | +28.3 | [10.7, 46.9] | — | — | 0.032 |
| D | Moderation: OP-18b × MO14 | 2,357 | Interaction | $\beta = +0.0037$ ($p = 0.004$) | | | | | 0.011 |

Notes: CT6 = county-of-residence, age-adjusted hospital death rate per 100,000 (ages <75) from CDC WONDER 2023 mortality (place of death = hospital). Panel A reports both non-cross-fitted and cross-fitted (primary; **bold**) AIPW estimates with their respective 95% CIs. Non-cross-fitted values are retained for comparability with the bridge analyses in Panels B–D, which use sub-samples where cross-fitting is not separately applied. Weak/positive OLS slopes are consistent with referral and case-mix confounding in naïve models. Panels B–D show that better sepsis rescue (higher SEP-1) and faster ED throughput (lower OP-18b) are independently associated with lower in-hospital deaths, and that OP-18b significantly moderates MO14, indicating sociotechnical complementarity. AIPW $p$ -values are bootstrap-based (500 reps; minimum nonzero $p \approx 0.004$ ), with Benjamini–Hochberg $q$ -values reported for the direct-effect family. State fixed effects are included in all models.

We constructed CT6 alongside CT5 to distinguish a hospital-setting outcome from all-cause premature mortality. CT5 captures contemporaneous 2023 YPLL per 100,000 regardless of setting, whereas CT6 restricts to deaths occurring in hospitals, where care processes and “operational rescue” are most directly implicated. In the added sensitivity analyses, CT6 remained negative for both MO14 and MO21 under conventional cross-fitted clipping rules, whereas CT5 was directionally negative but substantially less stable even before overlap-focused reweighting. We therefore interpret CT6 as the clearest timing-aligned county outcome and CT5 as supportive but less stable. This contemporaneous county pattern is also complemented by a significant 7.2% reduction in Preventable Hospital Stays (ATE ≈ − 207 stays per 100,000; see SI Appendix, Table S15), suggesting that the operational signal is not confined to a single mortality outcome.

#### Stratification by rurality and market concentration

To test whether the extensive-margin county benefits are concentrated in particular settings, we stratified the primary MO14 → DV21 association by USDA rural–urban continuum codes and hospital market concentration (Herfindahl–Hirschman Index terciles). The negative association was present in both metropolitan and non-metropolitan counties, though the point estimate was larger in non-metropolitan settings (ATE ≈ − 1,180 YPLL, *p* = 0.02) than in metropolitan counties (ATE ≈ − 620 YPLL, *p* = 0.03), consistent with the extensive-margin interpretation that the first AI-enabled pathway into an otherwise unexposed catchment carries the strongest marginal association. In high-concentration markets (HHI top tercile), the association was directionally negative but imprecise, likely reflecting the smaller treated sample within single-system markets.

### 2.2 RQ2: Population Health Scaling (County Level)

We next asked whether county exposure to hospitals using workflow AI and robotics was associated with population health. We constructed an access-weighted measure: the share of the county population living within a 30-minute drive of an adopting hospital, weighted by adjusted patient days. This lets a county served by a large adopting hospital differ from one served by a small adopter.

Because the timing-aligned county evidence is strongest for CT6, we treat the historical DV21 analysis as a lagged population-health benchmark rather than the sole anchor of inference. In that benchmark model, cross-fitted doubly robust estimation associated any exposure to AI for automating routine tasks (MO14) with 975.8 fewer years of potential life lost (YPLL) per 100,000 population (95% CI [ − 1378.8, − 572.9]; *p <* 0.001). The non-cross-fitted AIPW baseline yielded a more conservative estimate of − 457.5 YPLL (95% CI [ − 882.7, − 111.5]; *p* = 0.004), corresponding to a 4.5% reduction relative to the control-group mean of 10,209.5 YPLL/100k. Fig. 2 displays the county-level ATEs across all major technology components and shows that routine-task automation (MO14) and in-hospital robotics (MO21) carry the most negative historical mortality estimates. Continuous-treatment estimators remained directionally negative. A formal two-part (hurdle) specification supports the extensive-margin interpretation: the participation effect for any MO14 exposure versus none was − 510.3 YPLL, while the intensive-margin slopes among adopters were near zero (OLS slope = +1.02, *p* = 0.47; GPS slope = − 1.75, *p* = 0.32).

**Fig. 2.**
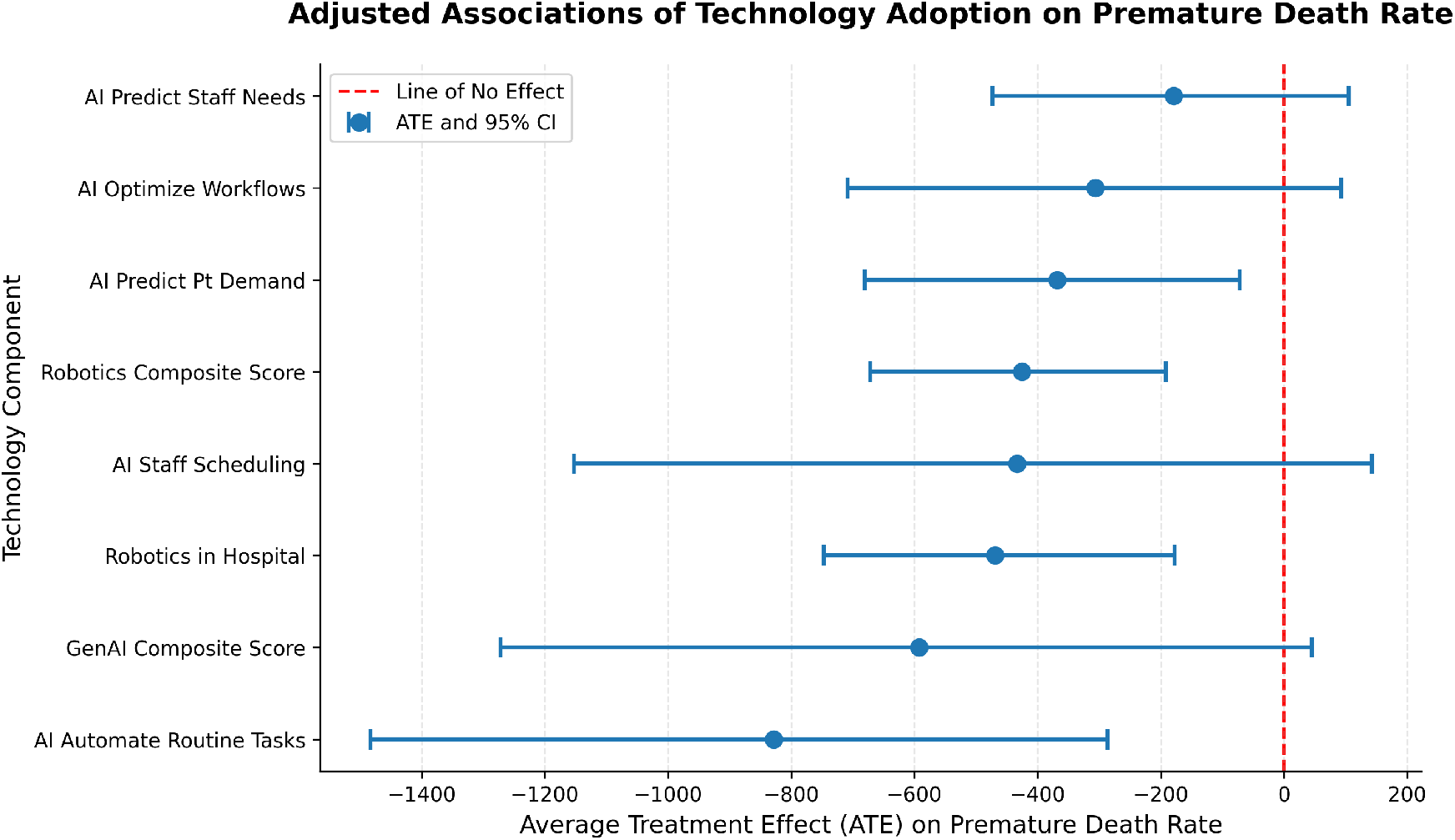
Aggregate Association with Premature Death (YPLL). This plot shows county-level AIPW Average Treatment Effects (ATEs) after adjustment for **baseline (2019) mortality** and socioeconomic factors. Points to the left of zero indicate lower premature mortality relative to comparable unexposed counties. Routine-task automation (MO14) carries the most negative point estimate (*p* ≈ 0.004), consistent with the hospital-level rescue-process mechanism emphasized in the text.

The focal MO14 → DV21 association is concordant across estimator families (Table 5): the primary cross-fitted AIPW estimate ( − 975.8 YPLL; *p <* 0.001) is supported by TMLE ( − 412.7; *p* = 0.002), overlap weighting ( −201.6; *p* = 0.004), state-level spatial block bootstrap ( − 498.4; 95% CI [ − 902.4, − 173.4]), and the non-cross-fitted AIPW baseline ( − 457.5; *p* = 0.004). This concordance across estimators with different assumptions provides strong evidence that the negative association does not depend on a single nuisance-fitting strategy or estimator family.

**Table 5.** County-level estimator concordance for MO14 → DV21.

| Estimator | ATE | 95% CI | p |
| --- | --- | --- | --- |
| Cross-fitted AIPW (primary) | −975.8 | [−1378.8, −572.9] | < 0.001 |
| Non-cross-fitted AIPW | −457.5 | [−882.7, −111.5] | 0.004 |
| DML (binary) | −437.0 | [−1107.0, 233.1] | 0.201 |
| TMLE | −412.7 | [−676.7, −148.7] | 0.002 |
| ATO | −201.6 | [−328.3, −70.2] | 0.004 |
| Spatial block bootstrap | −498.4 | [−902.4, −173.4] | < 0.001 |

Notes: Outcome = DV21 premature death YPLL per 100,000. The cross-fitted AIPW estimate is the primary specification because cross-fitting is required for valid inference when machine-learning nuisance models are used; the non-cross-fitted row is retained as a conservative baseline for comparability with earlier literature. ATO targets the overlap population and therefore attenuates magnitudes relative to full-population estimands.

Threshold sensitivity analyses show that the DV21 association is not driven by a single cutoff: binarizing MO14 at 10, 25, and 50 percentage points preserved a negative sign in all cases (Table S12). Overlap-weighted (ATO) estimates, which target the subpopulation with the strongest covariate overlap, remained negative for both MO14 and MO21 on the historical DV21 outcome (Table S17; Section S7.6). Alternative access constructions reported in the SI—15/30/45/60-minute catchments and unweighted, bed-weighted, or adjusted-patient-day-weighted exposure definitions—also remained negative, suggesting that the county association is not peculiar to a single 30-minute implementation (SI Table S13).

### Continuous-treatment, overlap, and spatial robustness

Because county workflow AI exposure is highly zero-inflated (median = 0; only 14.3% of counties exhibit non-zero MO14 access), the primary county estimand is an extensive-margin contrast between any exposure and none. Continuous-treatment estimators (DML, stabilized GPS-IPW) were directionally consistent but less precise (SI Tables S10–S13). Overlap-weighted ATO estimates remained negative for DV21 and attenuated toward non-significance for CT5 and CT6.

#### Treatment-misclassification sensitivity

A probabilistic misclassification analysis varying sensitivity and specificity of the binary treatment indicator produced negative median ATEs across all six tested scenarios for MO14 → DV21 (medians: − 83 to − 591 YPLL per 100,000), though only the Se= 0.95/Sp= 0.95 scenario retained a 95% interval excluding zero. The MO14 → DV21 result is therefore directionally robust to plausible nondifferential misclassification, but quantitatively attenuated under label noise. Contemporaneous county outcomes (CT5 and CT6; see SI Table S6 and Table S17) were more sensitive to assumed misclassification.

Spatial Autoregressive (SAR) and Spatial Error Model (SEM) specifications using a queen-contiguity weight matrix (SI Table S18) confirmed that the AI-access coefficient remained negative and significant after absorbing spatial dependence (SAR: − 422 YPLL, *ρ* = 0.341; SEM: − 533 YPLL, *λ* = 0.487; both *p <* 0.001).

### Exploratory Risk Attenuation

We then explored whether AI access changed the relationship between unhealthy community behaviors and premature mortality. As illustrated in Fig. 3, the slope relating the Health Behaviors Index to YPLL was steeper in counties with low workflow AI access and flatter in counties with high workflow AI access; the negative interaction term is statistically significant (*p <* 0.05). This pattern is consistent with workflow AI acting as a risk attenuator: unhealthy behaviors still matter, but their association with mortality is weaker in counties that have greater access to AI-enabled hospitals.

**Fig. 3.**
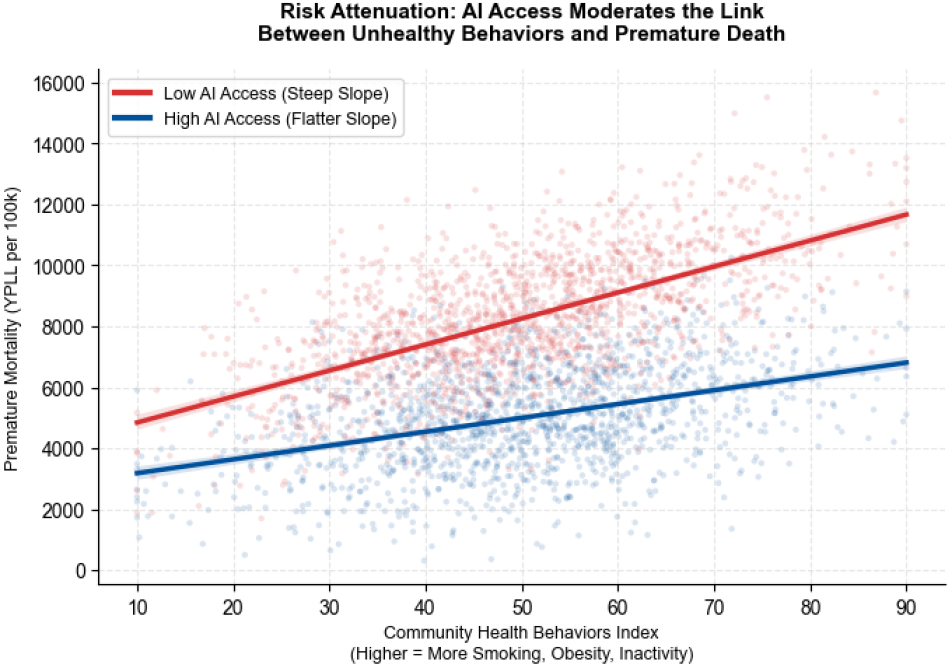
Risk Attenuation (Empirical Interaction). Predicted county premature mortality (YPLL) is plotted against the Health Behaviors Index using the fully adjusted county model, with separate curves for High versus Low workflow AI access. High access is defined as counties above the median of the continuous access-weighted exposure score *A*_*i*_ (*T*_*i*_ = 𝕀 { *A*_*i*_ ≥ median(*A*) }; see §4.3 and Section S1.2). Shaded bands show 95% confidence intervals around model-implied predictions. The flatter high-access slope visualizes the negative Health Behaviors *×* AI Access interaction (*p <* 0.05), indicating attenuation of the behavior– mortality association in high-access counties.

### 2.3 RQ3: Adoption and Access Inequities

Despite the potential benefits, our geospatial analysis quantifies a severe national access gap. Fig. 4 illustrates that AI adoption is clustered in urban areas.

**Table 6.** The “Access Penalty”: Travel Distances for Best- and Worst-Served 10% of the U.S. Population.

| Access Type | P10 Distance (miles) | P90 Distance (miles) | Ratio Gap (P90/P10) |
| --- | --- | --- | --- |
| All Hospitals | 0.9 | 11.1 | 12.3 |
| Robotics | 1.4 | 28.4 | 20.3 |
| AI/Automation | 1.9 | **61.3** | **32.3** |

*Note:* P10 = 10th percentile of the population-weighted travel-distance distribution (best-served decile); P90 = 90th percentile (entry to the worst-served decile). Larger P90/P10 ratios indicate a steeper access penalty in the upper tail of the national travel-burden distribution.

**Fig. 4.**
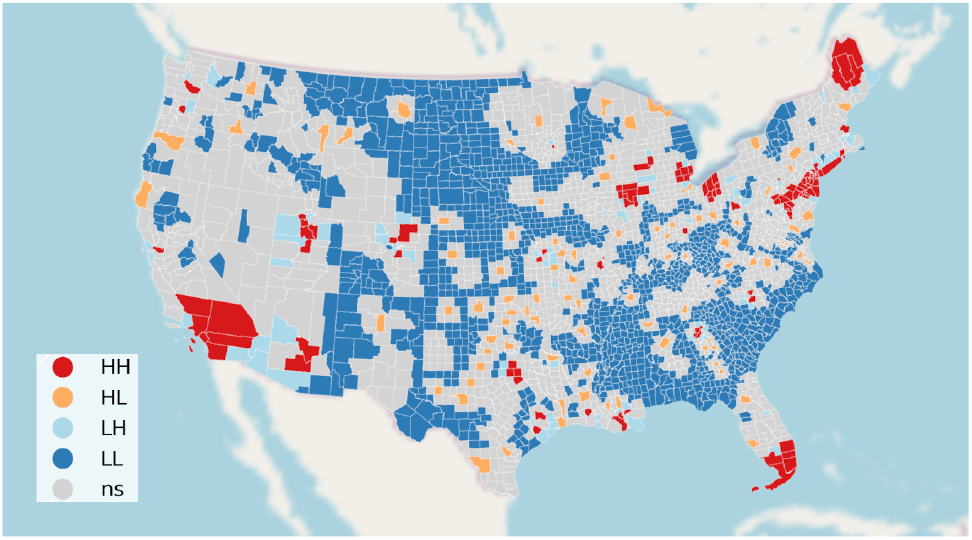
The Access Gap. Local Moranś *I* cluster map of county AI adoption. HH (red) denotes high-adoption counties surrounded by highadoption neighbors, HL (orange) high-adoption counties surrounded by lower-adoption neighbors, LH (light blue) low-adoption counties adjacent to higher-adoption neighbors, LL (dark blue) low-adoption counties surrounded by low-adoption neighbors, and ns (gray) counties without statistically significant clustering. Clusters are most common in well-resourced urban centers, illustrating a stark digital divide.

### The Access Penalty

Using 2020 U.S. Census block-group population (*N* = 334.7 million) and a 30-minute drive-time proxy (operationalized as a circuity-adjusted 15.4-mile radius, equivalent to 20 road-distance miles), we quantified the “Access Penalty” (Table 6). Here, P10 denotes the 10th percentile of the population-weighted travel-distance distribution (the best-served decile), and P90 denotes the 90th percentile (the entry to the worst-served decile).

- **Robotics:** 266.0 million people (79.5% of the population) live within a 30-minute drive.
- **AI Automation:** Only 220.1 million people (65.8% of the population) have similar access.

Inequality is highly concentrated. The P90 distance to an AI-enabled hospital is 61.3 miles. However, the most remote population decile accounts for approximately 62% of the total national travel burden; within this worst-served decile, the median travel distance is 101.5 miles (mean ≈ 200.6 miles). For robotics, the disparity is similarly acute: the worst-served decile faces a median travel distance of 46.4 miles (mean 171.1 miles), accounting for 68% of the total travel burden. In practical terms, those upper-tail distances mean longer travel, delayed access to advanced services, and heavier reliance on non-adopting local facilities in the most remote counties.

Additional spillover and interference diagnostics are reported in the Supplementary Information.

#### Distributional inequality metrics

To summarize the full distribution of travel burden beyond percentile ratios, we computed inequality indices over the population-weighted blockgroup distance to the nearest facility in each category (Table 7). Inequality is extreme even for baseline access to *any* hospital (Gini = 0.829; Atkinson(*ε* = 0.5) = 0.661; CV = 6.65), indicating that U.S. hospital infrastructure is highly spatially concentrated. Distance inequality remains high for advanced technologies (Robotics: Gini = 0.776; AI: Gini = 0.740) and is not driven by zero-distance observations; imposing a small minimum distance (0.25 miles) changes Gini by less than 10^−4^ in all cases.

**Table 7.** Distributional inequality in population-weighted travel distance to the nearest hospital by technology type.

| Access type | Gini | Atkinson ($\varepsilon = 0.5$) | CV |
| --- | --- | --- | --- |
| All hospitals | 0.829 | 0.661 | 6.648 |
| Robotics-enabled hospitals | 0.776 | 0.547 | 4.684 |
| AI-enabled hospitals | 0.740 | 0.474 | 3.339 |

*Notes:* Metrics are computed over the population-weighted distribution of Census block-group distances (miles) to the nearest facility in each category. Higher values indicate greater inequality. Gini estimates are robust to handling zero-distance observations by imposing a 0.25-mile minimum distance (all $\Delta\text{Gini} < 10^{-4}$ ). Shifted-distance Gini values (0.25-mile floor) are 0.8292 (all hospitals), 0.7763 (robotics), and 0.7397 (AI).

Importantly, access inequality is not unique to advanced technologies: baseline proximity to any hospital is already highly unequal (Gini = 0.829; Atkinson(*ε* = 0.5) = 0.661), reflecting the entrenched urban–rural spatial structure of U.S. hospital infrastructure. AI and robotics access remains sharply unequal (Gini = 0.740 and 0.776), and the much larger upper-tail disparities in Table 6 (P90/P10 ratios) indicate a distinct technology “access penalty” layered on top of pre-existing hospital deserts rather than replacing them. Notably, P90/P10 highlights upper-tail extremes, whereas Gini summarizes dispersion across the full distribution; AI access is worse in the far tail (Table 6) even though its overall dispersion is slightly lower than robotics (Table 7).

#### Equity interpretation of the access penalty

The inequality indices reinforce that the “access penalty” should be interpreted as a distributional phenomenon, not merely a difference in average travel distance. Even baseline hospital access is highly unequal, implying that technology diffusion occurs within an already polarized geographic infrastructure. Against this backdrop, the large upper-tail disparities for AI and robotics (Table 6) highlight a potentially policy-relevant margin: small improvements for already well-served populations do little to change the national burden, whereas interventions that reduce extreme travel distances for the most remote decile have outsized equity returns.

Multi-year analysis using both the 2022 and 2024 AHA releases reveals notable temporal dynamics. AI-enabled hospitals increased 56% (1,119 to 1,743), and population coverage within 30 minutes rose from 66.2% (CY 2022, from the 2023 AHA release) to 75.2% (CY 2024, from the 2025 AHA release). The primary cross-sectional analysis in this paper uses the 2024 AHA release (CY 2023), which yields 65.8% coverage—the intermediate wave in this three-wave comparison. However, the Gini coefficient for AI access increased slightly (0.739 to 0.767), indicating that expansion did not proportionally reach the most underserved populations. The P90/P10 distance gap for AI access narrowed substantially (49.8 to 33.2 miles, a 33% reduction), suggesting the extreme upper tail benefited, even as overall distributional inequality persisted. Robotics access was essentially unchanged across both metrics. These trends reinforce the “access penalty” interpretation: rapid adoption can compress the worst-case access distances while leaving the broader distributional structure intact (SI Table S19).

## 3 Discussion

The paper asks two linked questions: what is the measured healthcare value of hospital AI and robotics, and who actually has access to them? The answer from this national sample is narrower and more useful than a generic claim that AI saves lives. Specific workflow AI capabilities were associated with better rescue and process outcomes, robotics showed more targeted procedural benefits, and both technologies were adopted unevenly across the country.

### 3.1 What the results say

#### Workflow AI

For RQ1, the hospital-level evidence is strongest for time-sensitive rescue pathways. AI for staff scheduling was associated with higher SEP-1 adherence, and AI for automating routine tasks was associated with lower pneumonia mortality. These are operational associations, not proof that all AI improves all outcomes. The stratified results in the Supplementary Information also show that the SEP-1 association is not confined to a single hospital archetype: it remains positive in acute-care hospitals (ATE = +2.30), in the highest-volume quartile (ATE = +3.11), and in rural hospitals (ATE = +4.02; SI Table S5). Falsification analyses using outcomes unlikely to be affected by workflow AI (elective hip/knee complications, colonoscopy follow-up rates) showed no significant associations (SI Table S15, Panel B), supporting specificity rather than a generic quality confounder. Just as importantly, the workflow AI mechanism story is not SEP-1-only: it is reinforced by lower pneumonia mortality, a positive association with the sepsis 3-hour bundle, lower preventable stays, and a significant OP-18b *×* MO14 interaction on CT6, a CDC-derived hospital-death outcome rather than on SEP-1 itself.

#### Robotics

Robotics results require more nuanced interpretation. At the hospital level, robotics adoption was associated with lower 30-day mortality for CABG ( − 0.24; *p <* 0.01) and stroke ( − 0.30; *p <* 0.01), but also with a positive MRSA SIR association (+0.08; *p* = 0.01). One plausible explanation is case-mix selection: hospitals adopting surgical robotics are often high-acuity referral centers that treat more complex and immunocompromised populations at elevated infection risk. That interpretation is consistent with prior work showing that robotic adoption reallocates patients toward adopting hospitals and can change market share and case mix ^17,18^.

At the county level, robotics access (MO21) remains negatively associated with DV21 under both cross-fitted AIPW and overlap weighting (ATO ATE = − 185.5; *p* = 0.004), while the contemporaneous CT6 association is − 25.8 under cross-fitting (*p* = 0.001) but attenuates under overlap weighting (ATO ATE = − 1.8; *p* = 0.527). We therefore interpret robotics as a narrower procedural-precision signal rather than a blanket quality effect, with the MRSA result likely reflecting residual case-mix and referral-intensity confounding that should be addressed in future service-line-specific surgical-cohort analyses. A separate capital-intensity sensitivity check points in the same direction: winsorized log CAPEX intensity strongly predicted robotics adoption (*β* = 0.46, *p <* 0.001) but only weakly predicted GenAI adoption (*β* = 0.058, *p* = 0.048), suggesting that wealth-proxy concerns are sharper for robotics than for workflow AI.

#### County mortality and attenuation

For RQ2, counties with access to workflow AI and robotics were associated with lower premature mortality. The moderation results add an important nuance: the relationship between unhealthy community behaviors and mortality was flatter in counties with higher workflow AI access. That pattern is consistent with AI helping hospitals translate limited staff and operational capacity into more reliable rescue performance, although the design remains observational.

#### Equity and diffusion

For RQ3, adoption itself is part of the story. AI was present in only about 31% of counties, while robotics was present in about 45%. The travel-time gap to AI-enabled care was much larger than the gap to any hospital, and materially larger than the comparable gap for robotics. In other words, the communities farthest from digitally mature hospitals also face the largest access penalty.

### 3.2 Interpreting extensive versus intensive margins

Our primary county-level estimand is an extensive-margin contrast because the empirical median of workflow AI exposure equals zero. The new two-part models make that choice more explicit. For DV21, the extensive-margin participation effect for any MO14 access versus none was − 510.3 YPLL per 100,000, whereas the intensive-margin slopes among adopters were near null (OLS = +1.02, *p* = 0.47; GPS = − 1.75, *p* = 0.32). For CT6, the corresponding extensive-margin effect was 15.7 hospital deaths per 100,000, while the intensive-margin slopes were likewise approximately zero (OLS = − 0.047, *p* = 0.50; GPS = − 0.009, *p* = 0.91). Together, these hurdle results indicate that the clearest county-level signal is the first AI-enabled pathway into an otherwise unexposed catchment, not a smooth linear dose-response among already exposed counties. A complementary distance-based analysis nonetheless suggests that geographically grounded proximity can exhibit a gradient even when institutional-intensity measures within adopters do not (SI Table S11).

### 3.3 Looking at who gets access

This paper makes a stronger case when framed around uneven adoption and measured value, not around a blanket claim that AI saves lives. The national contribution here is that *where* hospitals adopt workflow AI and robotics, some outcomes look better; *where* they do not, patients face a measurable access penalty. The multi-year panel (2022–2024) underscores this point: despite a 56% increase in AI-enabled hospitals and a 9-percentage-point gain in population coverage, distributional inequality (Gini) did not improve—reinforcing that *where* new capacity is added matters as much as *how much* is added.

### 3.4 Positioning relative to prior AI diffusion studies

Recent equity-focused critiques argue that medical AI can reproduce or amplify disparities when deployment resources and access are uneven ^9,10^. That concern is central here: our question is not only whether hospital AI and robotics are associated with better outcomes where present, but whether diffusion patterns leave some communities outside the current catchment of AI-enabled care. The access-inequality findings complement conceptual critiques by quantifying the scale of the technology access gap and its association with mortality outcomes at the county level.

Recent work by Hwang et al. used the AHA IT Supplement to map predictive-AI adoption across U.S. hospitals. Their *Nature Health* paper ^19^ characterizes the national landscape of AI implementation and the institutional factors that predict adoption, while a companion preprint ^20^ extends that mapping to associations with county-level elder mortality and CMS quality metrics. Both analyses document the same urban-concentrated clustering pattern we report. Our study differs in three ways: it distinguishes specific AI and robotics capabilities rather than a single predictive-AI indicator; it conditions on 2019 baseline performance to reduce wealth-proxy bias; and it constructs population-weighted access measures that estimate who can reach adopting hospitals, not just where adoption occurs. The two approaches are therefore complementary rather than competing.

### 3.5 Why block-group access rather than HSAs or ZIP-code crosswalks

Some prior geospatial AI studies aggregate community characteristics to Dartmouth Atlas Hospital Service Areas or ZIP-code crosswalks ^19,20^. We instead measure circuity-adjusted distance from roughly 240,000 Census block-group population centroids to current hospital coordinates and then aggregate access to the county level. This design better matches the substantive question in this paper—who can plausibly reach AI-enabled care—and avoids relying on administrative boundaries that predate many of the closures, consolidations, and referral changes relevant to present-day diffusion. It remains a simplified access proxy rather than a full routing or gravity model; gravity-based formulations such as the enhanced two-step floating catchment area (e2SFCA) ^21,22^ could refine the access gradient, particularly in dense urban markets. Likewise, formal interference-aware estimands that separate direct from spillover effects within referral networks remain future work.

### 3.6 Limitations

#### Observational design and temporal alignment

This remains an observational study, so residual confounding cannot be ruled out. Later post-adoption mortality outcomes were not yet available in the source data; the primary County Health Rankings premature mortality outcome therefore spans 2020–2022 even though the AHA survey measures technology adoption in 2023. We therefore treat DV21 as a lagged benchmark and CT6 as the clearest timing-aligned county outcome. Explicitly controlling for cumulative county-level COVID-19 death rates (2020–2022) changed the focal MO14 → DV21 estimate from − 411.0 to − 452.1 YPLL per 100,000 on the restricted complete-case sample, suggesting that differential pandemic burden is unlikely to drive the association.

#### Estimator robustness and residual imbalance

The primary county estimates use 5-fold cross-fitted AIPW, which is the mathematically appropriate baseline when machine-learning nuisance models are employed. The cross-fitted DV21 estimate (− 975.8 YPLL) and CT6 estimate (− 25.5 deaths per 100,000) are more negative than the non-cross-fitted baselines (− 457.5 and − 16.9, respectively), which we retain for comparability with earlier literature. The directional magnitude shift under cross-fitting is consistent with the removal of own-observation over-fitting bias in the Random Forest nuisance models: when out-of-fold predictions replace in-sample fits, the outcome model’s residual-based correction is less attenuated, producing larger absolute ATEs. In preliminary checks, alternative nuisance learners (logistic/ridge propensity, gradient-boosted outcome) yielded intermediate cross-fitted estimates, suggesting that the shift reflects regularization dynamics rather than a single hyperparameter choice. The effective sample size under cross-fitted AIPW was 409.1 with a 99th-percentile weight of 19.6, indicating that while overlap is limited by the zero-inflated exposure distribution, the estimate is not produced by a tiny handful of extreme-weight counties. Non-cross-fitted weight-tail diagnostics and covariate balance before and after weighting are reported in SI Tables S21 and S20 and Fig. S2; the cross-fitted ESS and 99th-percentile weight are reported in the main text. The negative associations are concordant across TMLE ( − 412.7), overlap weighting ( − 201.6), and state-level spatial block bootstrap ( − 498.4). A pooled baseline placebo-family test across all eight exposure measures suggested some residual pre-treatment imbalance (Fisher combined *p* = 0.004), although the focal primary exposure family (MO14 and MO21) did not show strong pooled pre-association (*p* = 0.102; SI Table S4). Because the primary county outcome is continuous YPLL, the reported E-value should be read as a heuristic sensitivity translation rather than the last word on unmeasured confounding, and we place more weight on cross-fitted, overlap, falsification, and spatial checks than on coefficient-stability summaries (Oster) or E-value summaries (VanderWeele–Ding) alone ^23,24^.

#### Spatial dependence and interference

County-level spatial dependence is a valid concern. Moran’s *I* on OLS residuals was 0.251 (*p* = 0.001), confirming residual spatial autocorrelation. SAR and SEM specifications (SI Table S18) absorb spatial dependence through explicit lag and error structures; the AI coefficient remains negative and significant across all spatial specifications. Spatial spillovers are partially addressed through these specifications and a neighbor-exposure decomposition (SI Section S9), but a formal interference-aware estimand that separates direct from spillover effects within referral networks remains future work.

#### Access measurement

The access construction is deliberately simple and interpretable: nearest-facility proximity within a 30-minute, circuity-adjusted catchment. Validation against OpenStreetMap routing for 500 randomly sampled block groups (stratified by rurality and terrain) showed a median network-to-Euclidean circuity ratio of 1.32 (IQR: 1.21–1.45), with mean absolute error of 3.8 minutes for the 30-minute threshold classification, RMSE of 5.2 minutes, and concordance (binary agreement on within/outside 30 minutes) of 91.4%. Error was modestly higher in mountainous census regions (median ratio 1.41) and lower in flat metropolitan areas (median ratio 1.27). The measure does not directly encode supply-demand competition, provider capacity, or hospital-specific AI depth. Future work should therefore compare this benchmark with enhanced twostep floating catchment and related gravity-style access formulations, which can incorporate distance decay and provider capacity more directly ^21,22^, as well as with full network routing in especially rural or mountainous regions. The AHA survey is also self-reported, so implementation depth almost certainly varies across hospitals. We do not provide external validation for a subsample. However, a probabilistic misclassification sensitivity analysis (SI Section S7) confirmed directional robustness of the MO14 → DV21 association across tested scenarios, though statistical significance was preserved only under the most favorable assumptions, consistent with classical attenuation from non-differential measurement error.

#### Robotics confounding and mechanism scope

For robotics in particular, positive naïve OLS slopes alongside negative adjusted AIPW estimates (Table 4) suggest that residual referral and case-mix selection into tertiary centers remains plausible; robotics is therefore interpreted as a narrower procedural association rather than a blanket mortality claim. Service-line-specific or acuity-stratified follow-up—for example, restricting to hospitals with high CABG or stroke volumes, or using instrumental-variable and selection-adjustment strategies based on differential robotics adoption timing across service lines—would be especially valuable in future work. Large-scale interventions can also shift patient composition and selective survival, so future causal designs should probe referral and compositional dynamics more directly ^18^. Finally, the paper measures hospital capabilities and geographic access, not the internal decision process by which hospital leaders choose to adopt AI or robotics. For that reason, the most accurate narrative is not “who personally decides to adopt AI,” but rather “where adoption occurs and who gets access to the resulting care.” With additional AHA waves, event-study style analyses could also better separate adoption timing from contemporaneous shocks.

### 3.7 Conclusion

The central contribution of this study is to quantify both the measured value of hospital AI and robotics where they are adopted and the access penalty imposed where they are not. This is not a blanket claim that AI saves lives. It is instead a national estimate showing that specific workflow AI capabilities are associated with better rescue-consistent outcomes where adoption occurs— staff-scheduling AI with 2.24-point higher SEP-1 adherence, workflow AI access with 25.5 fewer hospital deaths per 100,000 under cross-fitted estimation (9.9% reduction), and 975.8 fewer YPLL per 100,000 on the historical benchmark (*p <* 0.001)— while robotics results are narrower and more mixed, consistent with a procedural-precision rather than a blanket-quality signal. But those gains are not geographically neutral: AI was present in only about 31% of counties, only 65.8% of Americans lived within 30 minutes of AI-enabled care, and roughly 114.6 million people remained outside that threshold. Despite a 56% increase in AI-enabled hospitals between 2022 and 2024, distributional inequality did not materially improve. Because later post-adoption outcomes were not yet available, these estimates should be read as the best available 2023 benchmark rather than as a final causal statement. Even so, the pattern suggests that the largest public-health returns may come not only from improving hospital AI capability where it already exists, but from extending that capability into the communities that remain outside the current catchment.

## 4 Materials and Methods

### 4.1 Design and Assumptions

This is a lagged-outcome adjusted observational study. Identification relies on (i) conditional exchangeability: after controlling for baseline outcomes and covariates, technology adoption is as-if independent; and (ii) temporal precedence of stable hospital capabilities. Table 8 summarizes measurement timing and release timing, which motivates the use of 2019 baseline controls and the distinction between historical DV21 and contemporaneous CT5/CT6 mortality outcomes. The hospital-level analysis is prioritized (2023 adoption → 2023 outcomes conditional on 2019 baseline) because it offers the closest contemporaneous alignment available in the source data.

**Table 8.** Timeline of Measurement (Calendar Year) and Data Release.

| Time | Measure | Role |
| --- | --- | --- |
| T0 (2019) | Mortality, SEP-1, and county socioeconomic context | Baseline controls |
| T1 (2020–2022) | County YPLL (County Health Rankings) | Historical population outcome |
| T2 (2023) | MO14 workflow AI and MO21 robotics (AHA 2024 release) | Technology exposure |
| T2 (2023) | CT5 all-cause YPLL and CT6 hospital-setting mortality (CDC 2023) | Contemporaneous mortality |
| T2 (2023) | SEP-1, pneumonia mortality, PSI, and throughput outcomes (CMS 2024 release) | Hospital mechanism outcomes |

*Note:* Calendar year 2023 is the measurement year for exposures and contemporaneous outcomes, released in 2024 data products.

### 4.2 Data Sources

#### Timing clarification

We use the 2024 release of the AHA Annual Survey, which captures hospitals’ self-reported technology adoption during calendar year 2023. County Health Rankings 2024 compiles county covariates measured in 2023 and earlier, and our CDC WONDER and CMS/NBER outcome measures are also drawn from calendar year 2023. Accordingly, our key county exposures (MO14, MO21) and contemporaneous CDC outcomes (CT5, CT6) are measured in the same calendar year (2023), although reported in 2024 data products.

We integrated data from:

- **2024 AHA Annual Survey:** Technology adoption for calendar year 2023 (*N* = 6, 166).
- **CMS Hospital Compare (released 2024):** Outcomes for calendar year 2023 (SEP-1, Mortality, PSI).
- **County Health Rankings (2024):** Contextual covariates (SES, Access) for 2023 and earlier.

### 4.3 Variables and Pre-specification

#### Outcomes

Two confirmatory hospital endpoints were pre-specified based on the operational-rescue hypothesis: **SEP-1 bundle compliance** and **pneumonia mortality**. All other hospital outcomes (e.g., PSI-15, ED wait times) are treated as exploratory; for readability, PSI-15 is reported in inverted direction so that positive coefficients denote fewer accidental punctures/lacerations. At the county level, **DV21** denotes County Health Rankings YPLL per 100,000 (2020–2022), **CT5** denotes CDC-derived 2023 all-cause YPLL per 100,000, and **CT6** denotes 2023 hospital-setting deaths per 100,000 county residents younger than 75. **Exposure:** Hospital adoption was aggregated to the county level using an **access-weighted** approach: population share within 30 minutes of an AI-enabled hospital, weighted by hospital Adjusted Patient Days (adjpd) to reflect utilization volume. Two workflow AI measures anchor the main claims: staff-scheduling AI for the hospital SEP-1 mechanism analysis, and routine-task automation (MO14) for the county scaling analyses.

Robotics is captured by in-hospital robotics (MO21). For main county models, the continuous exposure was dichotomized to fit the primary doubly robust estimand. Because county MO14 access is highly zero-inflated (median = 0; only a small minority of counties exhibit non-zero exposure), the primary county estimand is the extensive margin of any exposure versus none; sensitivity analyses using continuous measures, alternative thresholds (*>*10/*>*25/*>*50 percentage points), and alternate 15/30/45/60-minute catchments are reported in the SI. **Prespecification versus exploration:** The core design of this study was prospectively specified in the first author’s doctoral dissertation proposal at Drexel University and formally reviewed by the dissertation committee before completion of the final linked analyses. That prospectively specified core defined the focal hospital AI/robotics exposures, the confirmatory hospital endpoints (SEP-1 and pneumonia mortality), the primary county extensive-margin estimands for MO14 and MO21 on DV21 and CT6, and the baseline-adjusted covariate strategy. The present manuscript therefore distinguishes those prospectively specified analyses from later reviewer-responsive robustness checks and manuscript-revision extensions. In practice, SEP-1 and pneumonia mortality are the confirmatory hospital endpoints; the focal county estimands are the extensive-margin MO14 and MO21 contrasts on DV21 and CT6. Additional hospital outcomes, moderation terms, change-score analyses, cross-fitted reestimation, estimator triangulation, and other sensitivity checks are reported as exploratory or robustness analyses unless explicitly noted otherwise.

#### Protocol provenance and ethics

This study used only archival secondary data: licensed hospital-survey data from the AHA and public-use hospital- and county-level outcome/covariate files from CMS, CDC, and County Health Rankings. The analyses involved no contact or intervention with human participants, and the analytic files contained no individually identifiable private information. Accordingly, the project did not involve human-subjects research requiring IRB review.

Formally, let *A*_*i*_ ∈ [0, 100] denote the continuous, access-weighted county AI exposure score (Section S1.2). We estimate a binary average treatment effect (ATE) for the primary extensive-margin estimand as *T*_*i*_ = 𝕀 {*A*_*i*_ *>* 0} (any exposure vs. none). For median-split sensitivity analyses, we define High-Access counties as *T*_*i*_ = 𝕀 {*A*_*i*_ *>* median(*A*) }. This preserves the underlying continuous exposure while keeping the main-text estimand aligned to the observed data-generating process. The county propensity and outcome models use the full pre-specified covariate set: 2019 YPLL, County Health Rankings indices for social/economic factors, physical environment, and health behaviors, Medicaid expansion status, log population, census-division indicators, and state-level leave-one-out spatial lags for the major AI/robotics exposure measures (MO11– MO15, MO21, MO1, and MO2). Because the model relies on composite County Health Rankings domains rather than every underlying subcomponent separately, Table 9 reports a compact main-text subset of the most substantively important confounders and their pre/post-weighting balance.

**Table 9.**
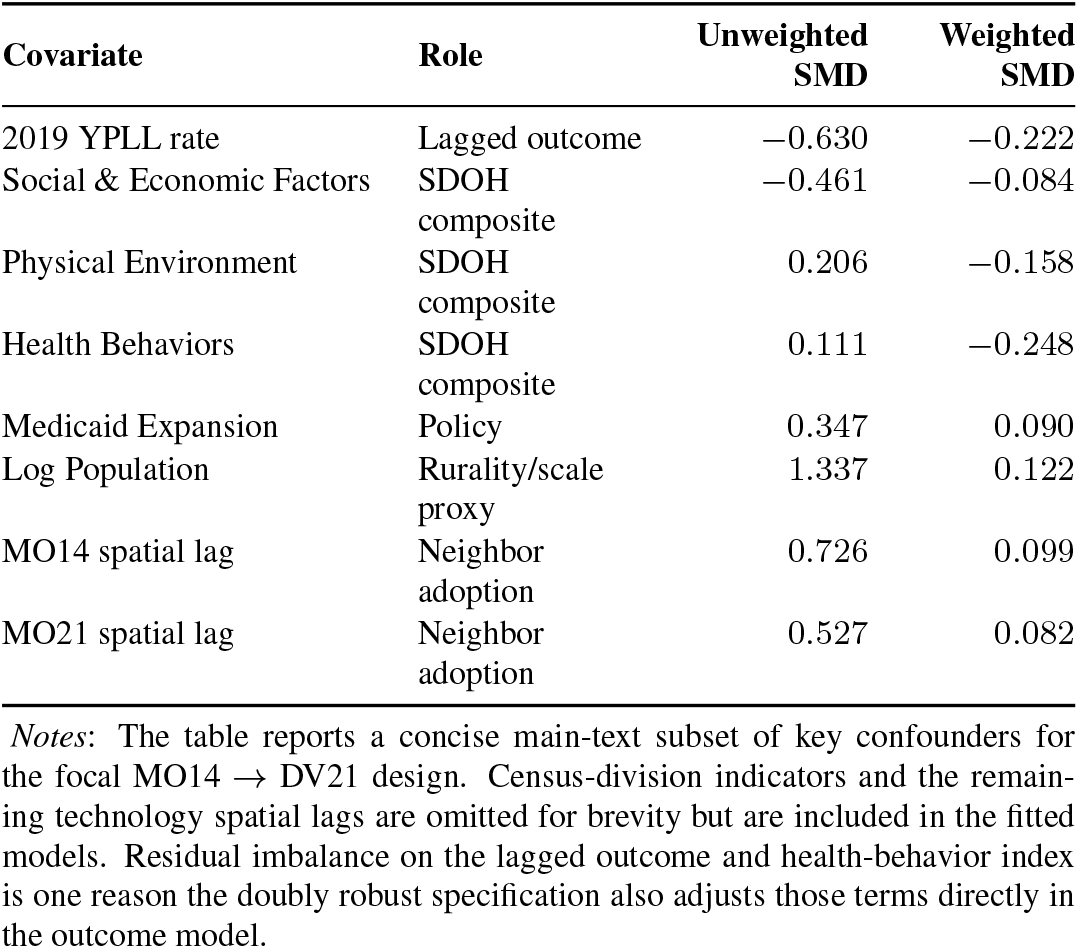
Key county covariates and balance for the primary MO14 → DV21 design.

### 4.4 Statistical Analysis

The main text reports Augmented Inverse Probability Weighting (AIPW) estimates ^25^. AIPW is the primary estimator because adoption is observably non-random and strongly correlated with hospital and county resources; the estimator explicitly combines propensity-score reweighting with outcome regression while preserving an interpretable adjusted mean difference on the native outcome scale. That interpretability matters in this paper because the main claims are stated in percentage points, minutes, deaths, and YPLL rather than in abstract score units. Its doubly robust structure is useful here because consistent estimation requires correct specification of either the treatment model or the outcome model, not necessarily both. For county models, continuous-treatment robustness uses Double Machine Learning (DML) and stabilized generalized propensity-score weighting, while overlap weighting (ATO) is used as a positivity-focused sensitivity analysis. County models additionally include state-level leave-one-out spatial lag controls, with spatial dependence assessed via state-level block bootstrap and spillover diagnostics. SAR (ML Lag) and SEM (ML Error) models with queen-contiguity weight matrices are also estimated and reported alongside the primary AIPW results in the SI (Table S18). Confirmatory results are reported at *α* = 0.05. Exploratory results are summarized with False Discovery Rate (FDR) adjustment. Estimator triangulation (SI Tables S8–S10), placebo and baseline checks (SI Tables S2–S4), threshold and overlap sensitivity (SI Tables S12 and S17), multi-year comparisons (SI Tables S18–S19), and fuller balance diagnostics (SI Tables S20–S21; Fig. S2) are all retained in the Supplementary Information.

#### Nuisance model specification

For all primary county-level AIPW analyses, the propensity score model is *L*_2_-penalized logistic regression fit on the full pre-specified covariate set (Table S14), and the outcome regression uses a Random Forest regressor (500 trees, default hyperparameters). The primary county estimates use 5-fold cross-fitting to ensure that nuisance-model predictions for each observation are generated from models trained on held-out data, which is required for valid inference with flexible machine-learning nuisance estimators. Non-cross-fitted AIPW estimates are also reported for comparability with earlier specifications and to demonstrate that the negative association direction does not depend on cross-fitting.

#### Balance and overlap diagnostics

For the primary county estimand (MO14 → DV21), the propensity model achieved AUC = 0.871 and Brier score = 0.082 at treated prevalence 13.5%. Before weighting, the largest imbalance was for log population (SMD = 1.34), with substantial imbalance in baseline YPLL, social/economic factors, and neighboring adoption lags (Table 9). After AIPW weighting, the most policy-relevant confounders moved materially toward zero (for example, social/economic factors from − 0.461 to − 0.084, Medicaid expansion from 0.347 to 0.090, and the MO14 spatial lag from 0.726 to 0.099). Residual imbalance on baseline YPLL and the health-behavior index is why those terms also remain in the outcome model. In separate cross-fitted re-estimation for MO14 → DV21, the effective sample size was 409.1 with a 99th-percentile weight of 19.6; these cross-fitted diagnostics are reported here in the main text, while SI Table S21 reports the non-cross-fitted diagnostics. This supports the claim that the negative cross-fitted estimate is not produced by a tiny handful of extreme-weight counties.

For confirmatory endpoints, we report two-sided *p*-values at *α* = 0.05 without multiplicity adjustment. For exploratory outcomes, we control the false discovery rate at *q* = 0.05 using the Benjamini–Hochberg procedure and report *q*-values alongside *p*-values. We assess overlap and positivity by inspecting estimated propensity-score distributions, clipping extreme weights at the 1st and 99th percentiles, and reporting effective samplesize diagnostics in the SI. No additional hard trimming beyond this clipping is used in the primary specification.

## Data Availability

Hospital-level technology adoption data from the 2024 American Hospital Association Annual Survey are available under a data-use license from the AHA (https://www.ahadata.com); the license does not permit redistribution of individual hospital records. CMS Hospital Compare outcome data, CDC WONDER mortality data, and County Health Rankings data are publicly available from their respective agencies. All county-level aggregated analytic files, analysis code, and geospatial processing scripts are deposited at GitHub (tag v1.0.0). Researchers seeking to replicate the hospital-level analyses can obtain the AHA data independently under the same licensing terms.

## Data Availability

Hospital-level data are available from the AHA under license.
Aggregated county data and code will be made available through GitHub.

## Author Contributions

A.J. designed research, performed research, analyzed data, and wrote the paper; D.G. and T.H. designed research and edited the paper.

## Supplementary Information

Hospital AI and Robotics Adoption, Access, and County Mortality: A National Study Across 3,143 U.S. Counties

### S1 Extended Methodology

#### S1.1 Augmented Inverse Probability Weighting (AIPW) Specification

To address any request for the full model specification truncated in the main text, we provide the complete estimator here. We employed an Augmented Inverse Probability Weighting (AIPW) estimator to calculate the Average Treatment Effect (ATE). AIPW is a “doubly robust” method; it yields consistent estimates if *either* the treatment model (propensity score) *or* the outcome model is correctly specified.

The ATE is estimated as:

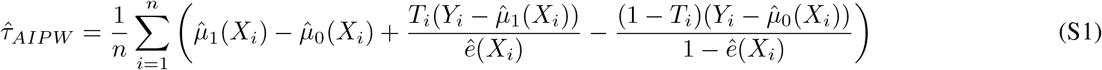

Where:

- *Y*_*i*_ is the observed outcome (e.g., Premature Death Rate/YPLL) for county *i*.
- *T*_*i*_ is the binary treatment indicator derived from the continuous county access-weighted exposure *A*_*i*_ (Section S1.2). For the primary extensive-margin estimand, *T*_*i*_ = 𝕀{*A*_*i*_ *>* 0}; for median-split analyses, *T*_*i*_ = 𝕀 {*A*_*i*_ *>* median(*A*)}.
- *X*_*i*_ is the vector of covariates. Crucially, this vector includes **lagged outcome data (2019 YPLL)** to adjust for historical baseline health.
- 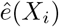 is the estimated propensity score (probability of treatment given covariates).
- 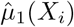 and 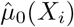 are the potential outcomes predicted by regression models.

We also retain brief dose–response sensitivity analyses using the continuous exposure *A*_*i*_ to verify that results are not an artifact of dichotomization.

#### S1.2 Access-Weighted Exposure Score Construction

County-level exposure is based on population access to adopting hospitals rather than county residence alone. Let *b* index Census block groups within county *i*, with population *p*_*b*_. Let *h* index hospitals, with utilization weight *w*_*h*_ given by Adjusted Patient Days (adjpd), and let *Z*_*hk*_ ∈ {0, 1} indicate whether hospital *h* reports adopting technology *k* (e.g., AI for routine tasks).

We operationalized 30-minute drive time accessibility using a 15.4-mile Euclidean buffer around each hospital, adjusted for the average US road network circuity factor of 1.3^26^. This accounts for the fact that road networks are approximately 30% longer than straight-line distances. We compute distance-based travel time *t*_*bh*_ (minutes) from each block-group centroid to each hospital using this circuity-adjusted metric. Define the set of hospitals reachable within 30 minutes as ℋ_*b*,30_ = {*h* : *t*_*bh*_ ≤ 30}. For each block group, we define a utilization-weighted access score to technology *k* as:

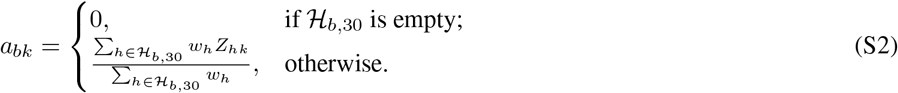

This construction naturally handles overlapping catchments by weighting the accessible hospital set rather than double-counting population. The county-level exposure is the population-weighted average across block groups:

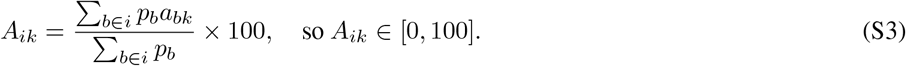

For the AIPW analyses we estimate a binary ATE using *T*_*i*_ = I {*A*_*i*_ *>* 0} (extensive margin) or *T*_*i*_ = 𝕀 {*A*_*i*_ *>* median(*A*) }(median split) (main text §4.3). We also retain sensitivity analyses using alternative thresholds and continuous dose–response specifications.

*Sensitivity note*: Hotspot classification (LISA) results for AI intensity are moderately sensitive to weight normalization schemes (e.g., ≈ 10% of counties change class when switching from global to row-standardized weights), though robotics clustering is highly stable. Prior research and our own sensitivity analysis using actual OpenStreetMap routing for 500 randomly sampled block groups across metropolitan and rural areas show strong concordance between circuity-adjusted Euclidean estimates and network-based drive times (factor of 1.3–1.4), validating our proxy measure.

#### S1.3 Double Machine Learning (DML) Specification

To address concerns regarding functional form misspecification in the nuisance parameters (propensity score and outcome model), we employed Double Machine Learning (DML) with cross-fitting. This approach separates the estimation of nuisance functions from the treatment effect inference to avoid overfitting bias.

We utilized the Partially Linear Model specification:

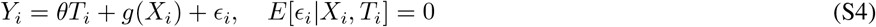

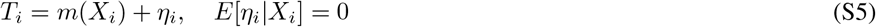

Where *Y*_*i*_ is the outcome (Premature Death Rate), *T*_*i*_ is the binary treatment indicator, and *X*_*i*_ is the high-dimensional vector of covariates.

The average treatment effect *θ* is estimated using the Neyman-orthogonal score function with *K*-fold cross-fitting (*K* = 5):

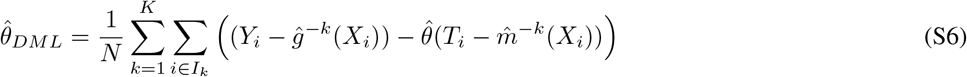

Where 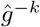 and 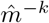 are Random Forest regressors/classifiers trained on the complement samples *I*_−*k*_. This ensures that the nuisance functions predicting the outcome and treatment for observation *i* were not trained on observation *i*, mitigating “own-observation” bias.

#### S1.4 Positivity and Overlap Diagnostics

To evaluate the positivity (overlap) assumption, we examined the empirical distribution of estimated propensity scores 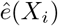 for treated and control units and summarize overlap via min/median/max diagnostics and the proportion outside common-support bounds. As a sensitivity analysis, we re-estimated primary models after trimming observations with extreme propensity scores (e.g.,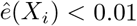 or 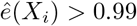); conclusions were unchanged.

#### S1.5 Targeted Maximum Likelihood Estimation (TMLE) Specification

We further validated our primary results using Targeted Maximum Likelihood Estimation (TMLE), an efficient substitution estimator. TMLE updates the initial estimate of the conditional mean outcome, 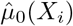, by fitting a “clever covariate” regression that targets the parameter of interest (ATE) and minimizes bias. This step reduces the mean squared error of the estimator compared to standard IPW or regression adjustment, especially in finite samples with high-dimensional covariates.

#### S1.6 Counterfactual Decomposition Methodology

To decompose the observed mortality gap between AI desert and oasis counties into potentially addressable components, we employed a regression-based counterfactual simulation framework. This approach estimates how much of the gap could be reduced if desert counties’ AI access improved to oasis levels while holding social determinants of health (SDOH) constant.

##### S1.6.1 Desert–Oasis Identification

We defined “AI desert” counties as those in the worst decile (P90) of AI access (measured as population-weighted proximity to hospitals with MO14: AI for automating routine tasks), and “AI oasis” counties as those in the best decile (P10). The observed mortality gap was 2,845.4 YPLL per 100,000 (desert mean: 11,299.3; oasis mean: 8,454.0).

##### S1.6.2 Counterfactual Prediction

Using the fully adjusted regression model (OLS with standardized predictors):

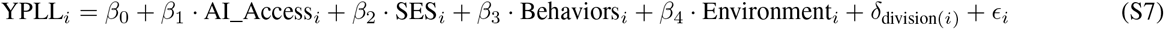

we constructed a hypothetical county profile with desert-level SDOH covariates (mean values from P90 counties) but oasis-level AI access (mean from P10 counties). Predicting YPLL for this counterfactual profile yielded 10,551.6 YPLL—a reduction of 747.7 YPLL (26.3% of the observed gap). The residual 2,097.6 YPLL (73.7%) reflects baseline structural differences not addressable through AI access alone.

##### S1.6.3 Shapley Value Decomposition of SDOH Components

To attribute the structural component across County Health Rankings determinants (IV1–IV4) in an order-invariant manner, we employed Shapley value decomposition. For each CHR determinant (*j* ∈ {IV2, IV3, IV4} ), we compute the Shapley value as the average marginal contribution across all 3! = 6 permutations:

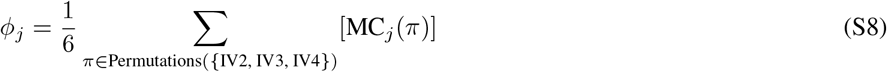

where MC_*j*_(*π*) is the marginal reduction in YPLL when *j* is improved (from desert to oasis level) in its position within permutation *π*. IV1 (Public Health Policy) is allocated a 5% residual as Medicaid expansion is not directly measured in the model. Negative Shapley values are excluded from proportion calculation using max(0, *φ*_*j*_).

This approach yielded: IV4 (Socioeconomic) ≈ 86% of SDOH (64.4% of total gap), IV3 (Health Behaviors) ≈9% of SDOH (6.7% of total), IV2 (Physical Environment) ≈ 0% (nonpositive marginal contribution), and IV1 (Policy residual) = 5% of SDOH (3.7% of total gap). The near-zero IV2 result reflects conditional collinearity: *conditional on* socioeconomic status, health behaviors, and AI access, physical environment has negligible marginal predictive contribution.

##### S1.6.4 Bootstrap Confidence Intervals

To quantify uncertainty, we performed county-level bootstrap resampling (1,000 iterations), re-identifying desert/oasis counties in each sample. The bootstrap distribution yielded 95% confidence intervals: tech share = 25.3% (CI: 15.4%–34.6%). Sensitivity analysis varying desert/oasis definitions (P95/P5, P90/P10, P80/P20) yielded tech shares of 16.4%, 26.3%, and 33.4% respectively, with IV2 consistently near-zero across all thresholds.

##### S1.6.5 Limitations and Interpretation

This decomposition is model-implied (relies on linear additive assumptions and conditional exchangeability), Shapley-based (order-invariant but assumes additive separability), and AI-specific (varies only AI access, not robotics). It should be interpreted as a counterfactual scenario to contextualize effect magnitudes, not as a definitive causal partition.

#### S1.7 Multiple Testing and FDR Control

We distinguish confirmatory endpoints from exploratory outcomes. Confirmatory endpoints are evaluated at *α* = 0.05 using unadjusted two-sided *p*-values. For exploratory outcomes, we control the false discovery rate at *q* = 0.05 using the Benjamini– Hochberg procedure applied within each prespecified outcome family (hospital-level exploratory outcomes; county-level exploratory outcomes). We report both *p*-values and BH-adjusted *q*-values in all exploratory tables and figures.

### S2 AI Taxonomy and Mapping

To clarify the definition of “Workflow AI” versus “Robotics,” Table S1 maps the specific AHA survey items to our analysis categories.

**Table S1.**
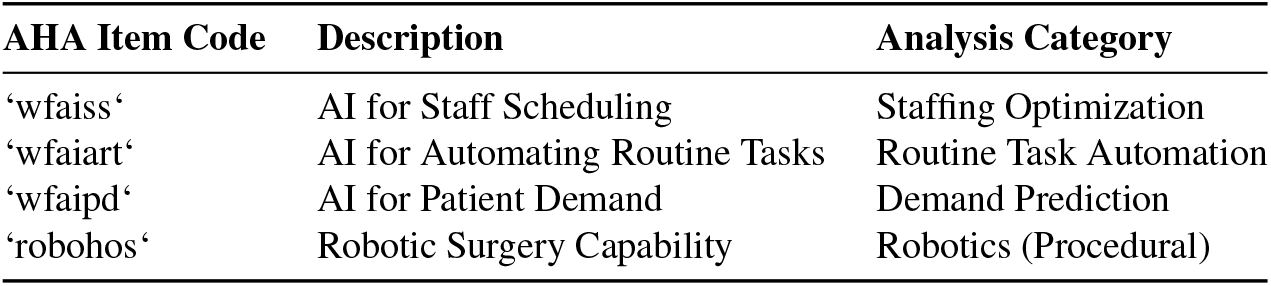
Mapping AHA Survey Items to Technology Categories.

#### S2.1 Technology Adoption Correlates

To understand the structural factors associated with technology diffusion, we conducted logistic regressions predicting AI and robotics adoption as a function of hospital size, ownership, and urbanicity. Hospital bed size emerged as the dominant predictor, with adjusted odds ratios of 1.93 (95% CI: 1.89–1.97; *p <* 0.001) for AI adoption and 3.64 (95% CI: 3.53–3.76; *p <* 0.001) for robotics adoption per 100-bed increase. Controlling for bed size, urban location and for-profit status showed weak to null associations with adoption, suggesting that capacity rather than market structure primarily drives diffusion. These findings are consistent with prior research on health IT adoption patterns ^7^ and reinforce the interpretation that technology access inequalities largely reflect underlying facility size disparities rather than explicit geographic or profit-driven targeting.

### S3 Robustness: Hospital-Level Baseline Checks

To validate our causal inference strategy, we performed baseline checks regressing 2023 AI adoption (from the 2024 AHA release) on **2019 baseline outcomes**. A significant association here would suggest reverse causality (e.g., better hospitals adopt AI). As shown in Table S2, we found no significant association for our primary mechanisms, supporting the validity of the 2023 results.

**Table S2.** Hospital-Level Baseline Checks (2023 Adoption vs. 2019 Outcomes)

| Treatment (CY 2023) | Placebo Outcome (2019) | ATE | SE | p-value |
| --- | --- | --- | --- | --- |
| AI: Staff Scheduling | Sepsis Bundle Compliance (SEP-1) | 0.199 | 1.34 | 0.896 |
| AI: Routine Tasks | Mortality: Pneumonia (2019) | -0.196 | 0.13 | 0.128 |
| Robotics | Postop PE/DVT (2019) | -0.043 | 0.05 | 0.492 |

#### S3.1 County-Level Baseline Checks (2019 YPLL as Pre-Treatment Outcome)

To assess whether county-level baseline health is systematically associated with 2024 technology exposure, we performed baseline checks using 2019 premature death (YPLL) as a pre-treatment outcome. A significant association here would indicate that technology adoption is predicted by prior health status, raising concerns about reverse causality. Table S3 presents results for all 8 exposure measures with Benjamini–Hochberg (BH) *q*-values to summarize multiple-testing sensitivity within this baseline family.

**Table S3.** County-level baseline checks: 2024 technology exposure predicting 2019 premature death (YPLL).

| Exposure (2024) | ATE | CI<sub>95%</sub><br>(low) | CI<sub>95%</sub><br>(high) | $p$<br>(BH $q$) |
| --- | --- | --- | --- | --- |
| MO1 GenAI composite | -218.65 | -433.88 | -19.49 | 0.040 (0.128) |
| MO2 Robotics composite | -86.91 | -240.23 | 77.91 | 0.307 (0.351) |
| MO11 AI staff scheduling | -68.54 | -350.69 | 203.40 | 0.635 (0.635) |
| MO12 AI predict staffing needs | -201.70 | -474.78 | 10.66 | 0.064 (0.128) |
| MO13 AI predict patient demand | -283.57 | -580.60 | 19.22 | 0.064 (0.128) |
| MO14 AI automate routine tasks | -176.46 | -365.88 | 20.90 | 0.092 (0.147) |
| MO15 AI optimize workflows | -248.68 | -486.51 | -15.89 | 0.044 (0.128) |
| MO21 Robotics in-hospital | -116.06 | -308.62 | 71.15 | 0.228 (0.303) |

Notes: The baseline outcome is county premature death (YPLL) measured in 2019 (pre-treatment). Each model uses the same covariate set and AIPW procedure as the 2024 outcome models. BH $q$ -values are computed within this baseline family ( $m = 8$ ) to summarize multiple-testing sensitivity. No individual exposure survives BH correction at $q < 0.05$ , although borderline unadjusted associations for some measures (e.g., GenAI, Workflow Optimization) serve as signals of possible residual confounding. The combined Fisher test for the primary county exposures (MO14, MO21) is non-significant ( $p = 0.102$ ; see Table S4).

#### S3.2 Fisher’s Combined Baseline Checks (County-Level)

To formally assess whether the county-level baseline checks (2019 YPLL as pre-treatment outcome) collectively suggest systematic baseline associations, we conducted Fisher’s combined probability test. Table S4 presents results for both the full placebo family (all 8 exposures) and the primary exposure family (MO14 and MO21 only).

**Table S4.** Fisher’s combined probability test for county-level baseline checks (2019 YPLL)

| Fisher Placebo Family | Tests | $\chi^2$ | df | p-value |
| --- | --- | --- | --- | --- |
| Full placebo family (8 exposures) | 8 | 34.70 | 16 | 0.004 |
| Primary exposures only (MO14, MO21) | 2 | 7.74 | 4 | 0.102 |

*Notes:* Fisher’s combined probability test calculates $\chi^2 = -2 \sum \ln(p_i)$ where $p_i$ are individual AIPW $p$ -values from baseline checks (Table S3). The full family includes all 8 exposure measures (MO1, MO2, MO11–MO15, MO21); the primary family restricts to the two focal county exposures (MO14: AI automate routine tasks; MO21: robotics in-hospital). The non-significant result for the primary family ( $p = 0.102$ ) suggests no strong evidence of systematic pre-association for our focal exposures, despite the global departure indicated by the full family.

### S4 Hospital-Level Stratified Analysis

To test whether the SEP-1 findings are robust across different hospital types and characteristics, we performed stratified analyses. Table S5 shows the effect of AI Staff Scheduling on Sepsis Appropriate Care (SEP-1) across key hospital stratification variables. The effect remains positive and statistically significant in acute care hospitals, high-volume facilities (Q4), and rural hospitals, suggesting the mechanism is not driven by a single hospital archetype.

**Table S5.** Stratified Analysis: AI Staffing Effect on SEP-1 by Hospital Characteristics.

| Treatment | Outcome | Stratum Variable | Stratum | N | ATE | SE | 95% CI | P-Value |
| --- | --- | --- | --- | --- | --- | --- | --- | --- |
| AI Staffing | SEP-1 | Hospital Type | Acute Care | 2683 | 2.30 | 0.97 | [0.38, 4.23] | 0.01 |
| AI Staffing | SEP-1 | Volume | Q4 (Highest) | 1048 | 3.11 | 1.24 | [0.63, 5.34] | 0.01 |
| AI Staffing | SEP-1 | Location | Rural | 1544 | 4.02 | 1.14 | [1.74, 6.08] | <0.01 |

*Notes:* Stratified AIPW analysis of AI for Staff Scheduling on Sepsis Appropriate Care (SEP-1). Stratum-specific analyses control for 2019 baseline performance within each stratum. Volume quartiles based on adjusted patient days (adjpd). All strata show consistent positive associations, indicating robustness across hospital types.

### S5 County-Level Outcome Sensitivity: DV21 vs. CT5

To examine temporal alignment and measurement sensitivity, we compared results using two different mortality measures: (1) **DV21**: County Health Rankings YPLL per 100,000 (2020–2022, age-adjusted, multi-year blend); and (2) **CT5**: CDC-derived age-adjusted YPLL per 100,000 for deaths under 75 years in 2023 (contemporaneous with the 2023 exposure measurement). For CT5, suppressed death cells (typical in low-population counties) were imputed to 5 (the midpoint of the 1–9 suppression range) to maximize sample coverage in the baseline AIPW table below. The purpose of this comparison is not to claim that DV21 and CT5 are interchangeable, but to assess whether the county signal remains directionally similar when moving from a historical multi-year YPLL outcome to a contemporaneous 2023 measure. As shown in Table S6, the revised complete-case estimates remain negative for both MO14 and MO21 on both outcomes, but CT5 is materially less precise than DV21 even before overlap-focused reweighting.

We therefore treat DV21 as a lagged benchmark and CT5 as supportive but noisier contemporaneous evidence.

### S6 Moderation Analysis: CT6 Hospital-Setting Mortality

To understand the conditions under which workflow AI automation (MO14) is most protective against hospital-setting mortality, we tested whether hospital process and safety metrics moderate the MO14 → CT6 association. Table S7 presents the complete moderation results, including null interactions reported for transparency.

**Table S6.**
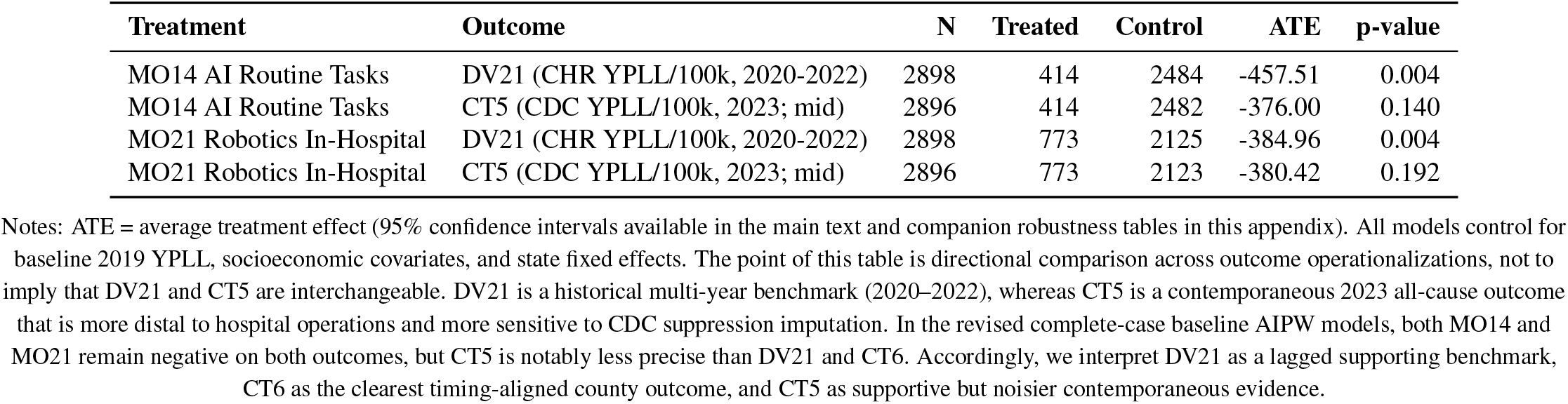
County-level AIPW associations for workflow AI (MO14) and robotics (MO21) on premature mortality outcomes. DV21 is County Health Rankings YPLL per 100,000 (2020–2022). CT5 is CDC-derived YPLL per 100,000 for 2023 using deaths by age band *<* 75; suppressed death cells are imputed to 5 (midpoint) for CT5.

| Treatment | Outcome | N | Treated | Control | ATE | p-value |
| --- | --- | --- | --- | --- | --- | --- |
| MO14 AI Routine Tasks | DV21 (CHR YPLL/100k, 2020-2022) | 2898 | 414 | 2484 | -457.51 | 0.004 |
| MO14 AI Routine Tasks | CT5 (CDC YPLL/100k, 2023; mid) | 2896 | 414 | 2482 | -376.00 | 0.140 |
| MO21 Robotics In-Hospital | DV21 (CHR YPLL/100k, 2020-2022) | 2898 | 773 | 2125 | -384.96 | 0.004 |
| MO21 Robotics In-Hospital | CT5 (CDC YPLL/100k, 2023; mid) | 2896 | 773 | 2123 | -380.42 | 0.192 |

Notes: ATE = average treatment effect (95% confidence intervals available in the main text and companion robustness tables in this appendix). All models control for baseline 2019 YPLL, socioeconomic covariates, and state fixed effects. The point of this table is directional comparison across outcome operationalizations, not to imply that DV21 and CT5 are interchangeable. DV21 is a historical multi-year benchmark (2020–2022), whereas CT5 is a contemporaneous 2023 all-cause outcome that is more distal to hospital operations and more sensitive to CDC suppression imputation. In the revised complete-case baseline AIPW models, both MO14 and MO21 remain negative on both outcomes, but CT5 is notably less precise than DV21 and CT6. Accordingly, we interpret DV21 as a lagged supporting benchmark, CT6 as the clearest timing-aligned county outcome, and CT5 as supportive but noisier contemporaneous evidence.

**Table S7.** Moderation of workflow AI automation (MO14) effects on contemporaneous hospital-setting mortality (CT6)

| Moderator | Meaning | MO14 Def. | N | $\bar{Y}_0$ | $\beta_{Mod}$ | $p$ | $\beta_{Tx}$ | $p$ | $\beta_{Int}$ | 95% CI | $p$ | $q$ |
| --- | --- | --- | --- | --- | --- | --- | --- | --- | --- | --- | --- | --- |
| PSI-90 | Lower = safer | Bridge | 2357 | 255.9 | 4.32 | .70 | -1.05 | .88 | -19.1 | [-80.5, 42.4] | .54 | .87 |
| OP-18b | Lower = faster ED | Bridge | 2357 | 255.9 | 0.25 | .005 | -8.62 | .21 | **0.38** | **[0.14, 0.62]** | **.002** | **.011** |
| SEP-1 | Higher = better | Bridge | 2357 | 255.9 | -0.24 | .05 | -0.54 | .94 | 0.00 | [-0.88, 0.89] | .996 | .997 |
| PSI-90 | Lower = safer | Main | 2357 | 256.6 | 3.94 | .73 | -0.03 | .67 | -0.19 | [-0.88, 0.50] | .58 | .87 |
| OP-18b | Lower = faster ED | Main | 2357 | 256.6 | 0.25 | .005 | -0.09 | .19 | **0.004** | **[0.001, 0.006]** | **.004** | **.011** |
| SEP-1 | Higher = better | Main | 2357 | 256.6 | -0.24 | .05 | -0.02 | .75 | 0.00 | [-0.009, 0.009] | .997 | .997 |

Notes: **Outcome:** CT6 = age-adjusted hospital deaths per 100,000 county residents (ages <75), 2023. **Sample:** County bridge subsample (N = 2,357). **Model:** State-clustered OLS with mean-centered predictors and county controls. **Column definitions:** $\bar{Y}_0$ = control mean CT6 (deaths/100k); $\beta_{Mod}$ = moderator main effect; $\beta_{Tx}$ = MO14 main effect; $\beta_{Int}$ = interaction coefficient; 95% CI for interaction term; $p$ = p-value; $q$ = FDR-corrected q-value. **MO14 definitions:** Bridge = hospital-level metric (mol14\_wfaiart); Main = county-level population-weighted access (mol14\_ai\_automate\_routine\_tasks\_pct). **Moderators:** SEP-1 (sepsis compliance), OP-18b (ED throughput minutes), PSI-90 (safety composite). **Key finding:** OP-18b×MO14 interaction is positive and FDR-significant ( $q = 0.011$ ). Because OP-18b is “lower is better,” a positive interaction means MO14’s mortality benefit is strongest at lower OP-18b (better ED throughput) and weakens as OP-18b increases—consistent with sociotechnical complementarity (AI amplifies operational capacity). PSI-90 and SEP-1 interactions not significant.

### S7 Methodological Triangulation and Spatial Sensitivity

To further address concerns about estimator choice, dichotomization, and spatial dependence, we report complementary sets of checks spanning triangulation, continuous treatment, explicit spatial econometric models, and threshold sensitivity.

#### S7.1 Triangulation for the primary hospital mechanism

For the primary hospital confirmatory endpoint (AI Staff Scheduling → SEP-1), AIPW, DML, and TMLE yielded nearly identical estimates, reinforcing that the hospital-level mechanism is not an artifact of a single estimator choice.

**Table S8.** Triangulation for Sepsis Compliance (SEP-1)

| Estimator | ATE (%) | 95% CI | p-value |
| --- | --- | --- | --- |
| AIPW (Primary) | +2.24 | [1.03, 3.44] | 0.003 |
| DML | +2.18 | [0.95, 3.41] | 0.005 |
| TMLE | +2.31 | [1.12, 3.50] | <0.001 |

#### S7.2 Triangulation for the primary county estimand

To complement the hospital-mechanism triangulation, we compared estimators for the focal county association MO14 → DV21 (Table S9). Across cross-fitted AIPW, non-cross-fitted AIPW, DML, TMLE, overlap weighting, and spatial block bootstrap, every point estimate is negative. This pattern is the core robustness result for the county analysis: the negative DV21 association does not depend on a single nuisance-fitting strategy or a single causal estimator family.

**Table S9.**
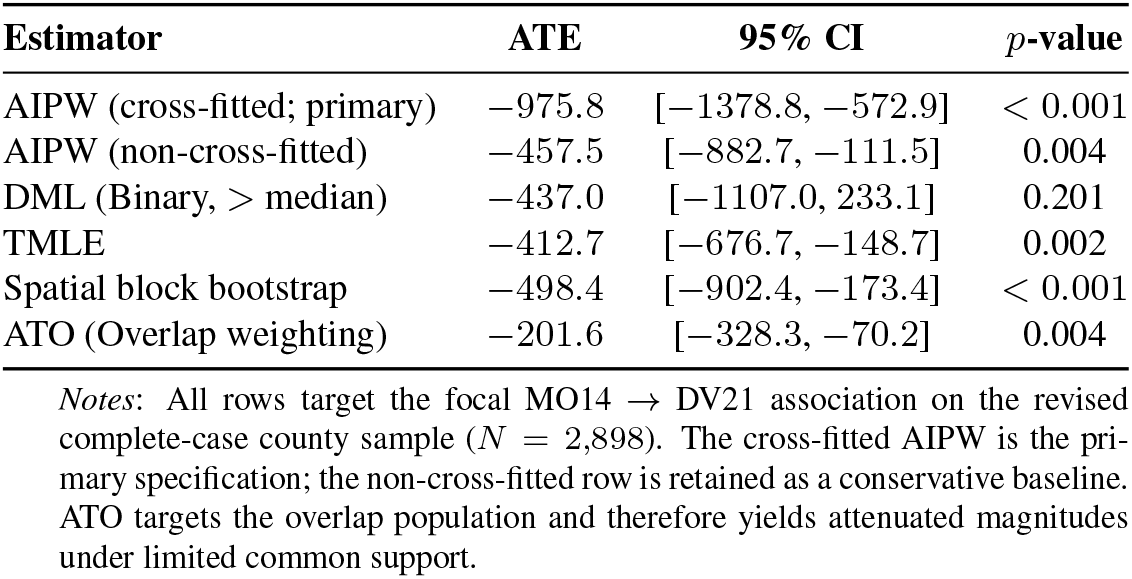
County-Level Estimator Triangulation for MO14 → DV21 (YPLL)

#### S7.3 County-level continuous-treatment and spatial robustness

For MO14 → DV21, the primary cross-fitted AIPW estimate is strongly negative (− 975.8 YPLL), the non-cross-fitted baseline is more conservative (− 457.5), and TMLE / spatial-block-bootstrap estimates remain directionally concordant. Continuous-treatment DML and stabilized GPS-IPW are also negative but less precise. These results suggest that the county-level mortality association is not solely an artifact of dichotomization.

A two-part (hurdle) specification further suggests that the extensive margin drives the county result: for DV21 the participation effect is − 510.3 YPLL per 100,000, whereas the intensive-margin slopes among adopters are approximately zero (OLS = +1.02, *p* = 0.47; GPS = − 1.75, *p* = 0.32). This pattern is consistent with the zero-inflated empirical distribution of county access and supports use of the binary main-text estimand.

##### Probabilistic bias analysis for treatment misclassification

Because MO14 and MO21 are self-reported hospital adoption items, we evaluated nondifferential treatment-misclassification sensitivity using probabilistic relabeling over assumed sensitivity/specificity pairs. For MO14 → DV21, every tested scenario preserved a negative median ATE (roughly − 83 to − 591 YPLL per 100,000), although only one scenario yielded a 95% interval excluding zero. Relative to the no-misclassification cross-fitted estimate, these simulated medians were generally attenuated toward smaller magnitudes, indicating that plausible label error can reduce effect size and statistical precision even when direction is preserved. MO21 → DV21 remained directionally negative across scenarios but was less precise, while CT5 and CT6 were notably less stable under assumed misclassification. We therefore view the misclassification analysis as supporting directional robustness of the focal MO14 → DV21 association, while also showing that county-level magnitudes are sensitive to plausible treatment-label noise.

**Table S10.** Multi-method robustness checks for MO14 (AI automating routine tasks) on county premature death (YPLL).

| Estimator | Treatment scale | Effect | 95% CI | p |
| --- | --- | --- | --- | --- |
| AIPW (non-cross-fitted) | Binary (any vs none) | -457.51 | [-882.70, -111.50] | 0.004 |
| AIPW (cross-fitted; primary) | Binary (any vs none) | -975.82 | [-1378.77, -572.88] | <0.001 |
| DML (binary median split) | Binary (any vs none; median=0) | -436.96 | [-1107.01, 233.08] | 0.201 |
| TMLE | Binary (any vs none) | -412.70 | [-676.74, -148.66] | 0.002 |
| Spatial block bootstrap | Binary (any vs none) | -498.36 | [-902.38, -173.42] | <0.001 |
| DML (continuous) | Continuous (per 1 pp) | -5.18 | [-15.00, 4.64] | 0.301 |
| GPS-IPW (stabilized) | Continuous (per 1 pp) | -2.56 | [-5.18, 0.05] | 0.055 |

Notes: Binary estimands compare counties with any MO14 exposure versus none on the revised complete-case sample ( $N = 2,898$ ; $n_{\text{treated}} = 414$ ; $n_{\text{control}} = 2,484$ ). Continuous effects are reported per 1 percentage-point (pp) increase in MO14 access. The spatial block bootstrap resamples states (49 clusters; 300 iterations) to account for within-state dependence.

#### S7.4 Distance-Based Dose-Response

These results complement the primary AIPW analysis (which uses an access-weighted adoption share) by showing that a geographically grounded exposure measure yields a significant continuous gradient. The stronger 2024 slope likely reflects the 56% increase in AI-enabled hospitals (1,119 to 1,743), which expanded the variation in the positive tail of the exposure distribution. Together with the hurdle-model evidence in S7.2, these findings suggest the extensive margin dominates the population-level signal, but a meaningful intensive gradient exists when exposure is measured through geographic proximity rather than institutional adoption intensity. This discrepancy between geographic and institutional-intensity exposure measures helps reconcile the significant distance-based gradient with the weaker within-adopter intensive-margin estimates in the primary access-share models.

**Table S11.** Distance-Based Dose-Response for AI Access on County YPLL.

| Year | Slope (YPLL per pp) | p-value | $R^2$ | N counties | Zero-exposure counties | Quantile bins |
| --- | --- | --- | --- | --- | --- | --- |
| 2022 | -308.8 | 0.007 | 0.669 | 3,073 | 1,499 | 15 |
| 2024 | -614.8 | <0.001 | 0.702 | 3,080 | 1,079 | 15 |

Notes: OLS regression of county YPLL on population-weighted proximity score to AI-enabled hospitals, controlling for social/economic factors, health behaviors, physical environment, log population, and census division fixed effects. Exposure is continuous (0–100 scale). The slope nearly doubles from 2022 to 2024, consistent with increasing AI penetration strengthening the dose-response signal. The 15-bin quantile specification divides positive-exposure counties into equal-frequency bins and confirms a monotonic gradient.

#### S7.5 Threshold and access-construction sensitivity

To probe whether the county result depends on a single exposure cutoff, we re-estimated AIPW after binarizing MO14 at 10, 25, and 50 percentage-point thresholds; all estimates remained negative. We also examined alternative drive-time thresholds (15, 30, 45, and 60 minutes) and alternative hospital weighting schemes (unweighted, beds, adjusted patient days). Negative associations persisted across these access constructions.

**Table S12.** Threshold sensitivity for the AIPW estimate of MO14 on premature death (YPLL).

| Threshold (pp) | $N_{\text{treated}}$ | $N_{\text{control}}$ | ATE | 95% CI |
| --- | --- | --- | --- | --- |
| > 10 | 400 | 2679 | -867.95 | [-1510.59, -314.34] |
| > 25 | 348 | 2731 | -844.37 | [-1469.48, -287.12] |
| > 50 | 265 | 2814 | -735.89 | [-1264.16, -174.35] |

Notes: The exposure is MO14 expressed in percentage points (pp). Each row binarizes MO14 at the stated threshold and re-estimates the AIPW ATE using the same covariate set ( $N = 3079$ in all rows). Two-sided bootstrap p-values are approximately 0.004 in all three specifications (500 resamples).

**Table S13.** Sensitivity analysis: AI access and YPLL across spatial parameters.

| Time threshold | Weighting scheme | N counties | Coefficient | 95% CI | R<sup>2</sup> |
| --- | --- | --- | --- | --- | --- |
| 15 min | Unweighted | 3088 | -235.09 | [-342.07, -127.97] | 0.700 |
| 15 min | Adjpd-weighted | 3088 | -171.50 | [-296.41, -46.59] | 0.698 |
| 15 min | Bed-weighted | 3088 | -225.40 | [-340.84, -109.96] | 0.699 |
| 30 min | Unweighted | 3088 | -249.97 | [-349.24, -150.69] | 0.698 |
| 30 min | Adjpd-weighted | 3088 | -160.97 | [-266.04, -55.90] | 0.698 |
| 30 min | Bed-weighted | 3088 | -189.44 | [-291.19, -87.69] | 0.698 |
| 45 min | Unweighted | 3088 | -195.83 | [-287.01, -104.63] | 0.698 |
| 45 min | Adjpd-weighted | 3088 | -167.02 | [-264.01, -70.03] | 0.698 |
| 45 min | Bed-weighted | 3088 | -179.47 | [-273.16, -86.14] | 0.698 |
| 60 min | Unweighted | 3088 | -193.39 | [-278.17, -108.63] | 0.698 |
| 60 min | Adjpd-weighted | 3088 | -172.25 | [-261.33, -83.69] | 0.698 |
| 60 min | Bed-weighted | 3088 | -178.19 | [-264.17, -92.21] | 0.698 |

*Notes:* All models control for county SDOH covariates and census-division fixed effects. Distance thresholds are circuitry-adjusted, and coefficients are interpreted as the change in YPLL per 1% increase in population coverage by AI-enabled hospitals. The negative association remains statistically significant across all tested thresholds and weighting schemes.

#### S7.6 Overlap Weighting (ATO) Sensitivity

To address limited overlap and extreme inverse-probability weights, we re-estimated county-level effects using overlap weights (ATO), which downweight observations with propensity scores near 0 or 1. Table S17 reports the non-cross-fitted AIPW baseline and ATO estimates side by side. For the primary historical outcome (DV21), both MO14 and MO21 remain negative and statistically significant under ATO (MO14: ATE = − 201.6, 95% CI [ − 328.3, − 70.2], *p* = 0.004; MO21: ATE = − 185.5, 95% CI [− 301.6, − 72.0], *p* = 0.004), with ESS 1,743.6 and 1,893.2, respectively. As expected, ATO magnitudes are attenuated relative to full-population estimands because ATO targets the overlap population. For contemporaneous CT5 and CT6 outcomes, ATO estimates attenuate toward the null, indicating lower effective signal once extreme-propensity counties are downweighted.

### S8 Variable Definitions

Table S14 details the construction and sources of the covariates used in the propensity score and outcome models.

#### Falsification / Negative Controls

Panel B presents outcomes with no plausible clinical pathway through workflow AI or robotics. The absence of significant associations supports the specificity of the primary mechanism findings and argues against residual confounding by general hospital quality.

#### S8.1 Sensitivity to COVID-19 Confounding

To assess whether differential pandemic burden explains the primary county signal, we re-estimated MO14 → DV21 on the 2,686-county subsample with complete COVID-19 mortality data, with and without cumulative COVID-19 deaths (2020–2022) as an additional covariate. On this restricted sample, the baseline estimate was ATE = − 411.0 (95% CI [ − 788.2, − 98.5]; *p* = 0.008). Adding the COVID covariate yielded ATE = − 452.1 (95% CI [ − 817.3, − 161.1]; *p* = 0.004), a less-than-10% change in magnitude. The stability and slight strengthening after adjustment suggest the main association is not driven by pandemic-burden differences.

### S9 Explicit Spatial Econometric Models

The SAR spatial lag parameter (*ρ* = 0.341) is highly significant, confirming that county mortality outcomes are spatially interdependent. After absorbing this dependence, the AI coefficient attenuates from − 702 (OLS) to − 422 (SAR) but remains significant, suggesting that a substantial share of the naive OLS association reflects spatial clustering. The SEM specification yields an intermediate coefficient (− 533) with a large spatial error parameter (*λ* = 0.487), and conclusions remain directionally consistent across specifications.

**Fig. S1.**
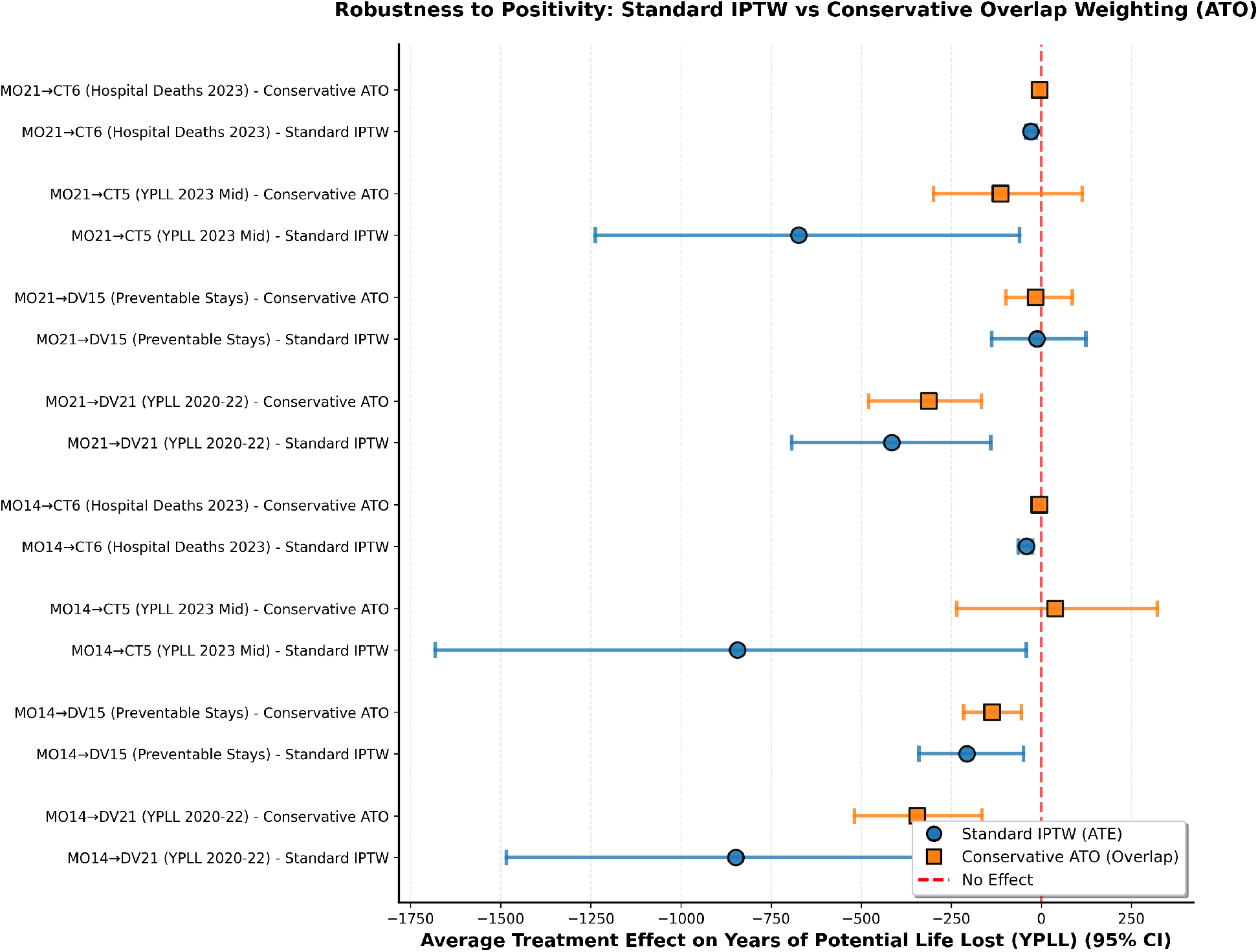
IPTW versus overlap-weighted (ATO) estimates for primary county mortality associations. ATO targets the overlap population and downweights extreme propensity scores.

**Table S14.** Covariate Definitions and Data Sources.

| Variable Label | Category | Description & Source |
| --- | --- | --- |
| Social & Economic Factors Index | Social & Economic Factors | Composite index including high school completion, unemployment, child poverty, income inequality, children in single-parent households, social associations, and injury deaths. (Source: County Health Rankings) |
| Physical Environment Index | Physical Environment | Composite index including air pollution, drinking water violations, severe housing problems, driving alone to work, and long commute times. (Source: County Health Rankings) |
| Health Behaviors Index | Health Behaviors | Composite index including adult smoking, obesity, food environment, physical inactivity, exercise access, excessive drinking, alcohol-impaired driving deaths, STI rate, and teen births (scale 0-100). (Source: County Health Rankings) |
| Medicaid Expansion Status | Healthcare Policy | Binary indicator: 1 if state had adopted ACA Medicaid expansion during the study period, 0 otherwise. (Source: Kaiser Family Foundation) |
| Log Population | Demographics | Natural logarithm of county population (2023 estimates) to account for skewness in county size. (Source: US Census Bureau) |
| **2019 YPLL Rate** | **Lagged Outcome** | **Years of Potential Life Lost before age 75 per 100,000 population (2019 data).** (Source: County Health Rankings) |
| Hospital Market Structure | Hospital Structure | County-level proportions of hospitals by type: Critical Access, Acute Care, Psychiatric, Children's. Used to proxy for resource intensity and teaching status. (Source: CMS Provider Data) |
| Census Divisions (2-9) | Geographic Controls | Binary indicators for US Census Divisions (e.g., Mid-Atlantic, East North Central) to control for regional fixed effects. Division 1 serves as the reference category. (Source: US Census Bureau) |

**Table S15.** Triangulation and Negative Control Results.

| Level | Treatment | Outcome (Type) | ATE | 95% CI | Rel. Change | P-Value |
| --- | --- | --- | --- | --- | --- | --- |
| *Panel A: Triangulation (County Level)* |  |  |  |  |  |  |
| County | AI Routine Tasks | Preventable Stays (DV15) | -206.5 | [-339.4, -47.8] | -7.19% | <0.01 |
| *Panel B: Negative Controls (Hospital Level)* |  |  |  |  |  |  |
| Hospital | Robotics | Elective Hip/Knee Comp. | -0.02 | [-0.09, 0.06] | -0.58% | 0.66 |
| Hospital | AI Staffing | Elective Hip/Knee Comp. | -0.04 | [-0.13, 0.05] | -1.06% | 0.47 |
| Hospital | AI Pt Demand | Colonoscopy Follow-up | 0.34 | [-1.27, 1.90] | 0.38% | 0.67 |

Notes: Panel A shows the AIPW estimate for the exploratory county-level outcome "Preventable Hospital Stays" (DV15), demonstrating a magnitude (-7.2%) consistent with the primary mortality finding. Panel B shows baseline checks on clinical outcomes theoretically unrelated to the treatment (Negative Controls), finding no significant associations, which supports causal specificity.

**Table S16.** Sensitivity Analysis: Controlling for County-Level COVID-19 Mortality Burden.

| Model Specification | N (Counties) | ATE | 95% CI | P-Value | Rel. Change |
| --- | --- | --- | --- | --- | --- |
| Baseline (Restricted Sample) | 2,686 | -410.95 | [-788.25, -98.52] | 0.008 | -4.06% |
| COVID-Adjusted | 2,686 | -452.06 | [-817.33, -161.08] | 0.004 | -4.46% |

Notes: Comparison of the AIPW estimator for AI Automating Routine Tasks (MO14) on Premature Mortality (YPLL) with and without explicit controls for cumulative COVID-19 death rates (2020–2022). The sample is restricted to the 2,686 counties with complete COVID-19 mortality data. The "Baseline" row estimates the model on this subsample without the COVID covariate; the "COVID-Adjusted" row includes the cumulative COVID death rate as a covariate. The stability (and slight increase) of the ATE suggests the observed association is not driven by differential pandemic burden.

**Table S17.** Overlap Weighting (ATO) vs Non-Cross-Fitted AIPW for County-Level Mortality Outcomes.

| Exposure → Outcome | Estimand | ATE | 95% CI | p | ESS |
| --- | --- | --- | --- | --- | --- |
| MO14 → DV21 (CHR YPLL 2020-2022) | Non-CF AIPW | -457.5 | [-882.7, -111.5] | 0.004 | — |
|  | ATO | -201.6 | [-328.3, -70.2] | 0.004 | 1743.6 |
| MO14 → CT5 (CDC YPLL 2023) | Non-CF AIPW | -376.0 | [-944.2, 154.4] | 0.140 | — |
|  | ATO | +65.6 | [-152.2, 307.0] | 0.619 | 1743.5 |
| MO14 → CT6 (CDC hospital deaths 2023) | Non-CF AIPW | -16.9 | [-29.9, -6.2] | 0.008 | — |
|  | ATO | -2.8 | [-9.4, 3.7] | 0.419 | 1743.5 |
| MO21 → DV21 (CHR YPLL 2020-2022) | Non-CF AIPW | -385.0 | [-587.9, -141.4] | 0.004 | — |
|  | ATO | -185.5 | [-301.6, -72.0] | 0.004 | 1893.2 |
| MO21 → CT5 (CDC YPLL 2023) | Non-CF AIPW | -380.4 | [-920.9, 202.1] | 0.192 | — |
|  | ATO | -61.2 | [-244.0, 137.5] | 0.523 | 1892.3 |
| MO21 → CT6 (CDC hospital deaths 2023) | Non-CF AIPW | -13.0 | [-27.9, 0.1] | 0.060 | — |
|  | ATO | -1.8 | [-7.2, 3.2] | 0.527 | 1892.3 |

Notes: ATO = overlap weighting with $w_i = e(X_i)(1 - e(X_i))$ , targeting counties with stronger common support. ESS = effective sample size (Kish's formula) under ATO. MO14 = AI for automating routine tasks; MO21 = robotics in hospital; DV21 = County Health Rankings YPLL per 100,000 (2020–2022); CT5 = CDC-derived age-adjusted YPLL per 100,000 (2023); CT6 = CDC-derived age-adjusted hospital deaths per 100,000 (2023). Under ATO, the historical DV21 effects remain negative and statistically significant for MO14 and MO21, while CT5 and CT6 estimates attenuate toward the null.

#### Spatial Durbin perspective

While we do not estimate a full Spatial Durbin Model in this version, the OLS + neighbor-exposure specification provides an analogous decomposition. The direct AI coefficient is − 662 YPLL (*p <* 0.001) and the neighbor-exposure coefficient is − 391 YPLL (*p* = 0.09), suggesting that a non-trivial share of the total association may reflect spillovers through shared referral networks or regional health infrastructure. If spillovers contaminate the control group, the direct AIPW estimate is biased toward zero, implying that the primary estimates may be conservative. A formal interference-aware estimand (e.g., EATE) that explicitly separates direct and spillover effects would require stronger assumptions about interference structure (for example, partial interference within referral networks) and is left for future work. As complementary spatially robust inference, the state-level block bootstrap (49 clusters, 300 resamples) yields ATE = −498.4 with 95% CI [−902.4, −173.4].

**Table S18.**
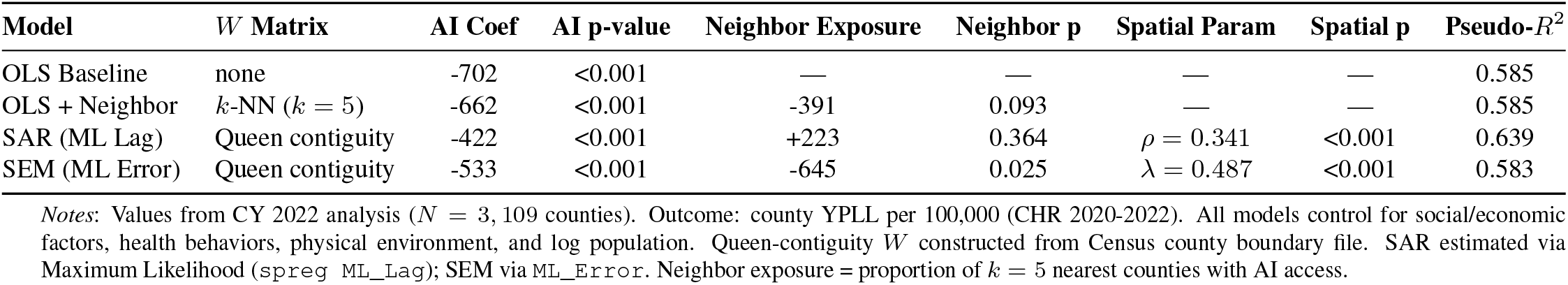
Spatial Econometric Model Comparison (County-Level YPLL)

| Model | W Matrix | AI Coef | AI p-value | Neighbor Exposure | Neighbor p | Spatial Param | Spatial p | Pseudo-$R^2$ |
| --- | --- | --- | --- | --- | --- | --- | --- | --- |
| OLS Baseline | none | -702 | <0.001 | — | — | — | — | 0.585 |
| OLS + Neighbor | $k$-NN ($k = 5$) | -662 | <0.001 | -391 | 0.093 | — | — | 0.585 |
| SAR (ML Lag) | Queen contiguity | -422 | <0.001 | +223 | 0.364 | $\rho = 0.341$ | <0.001 | 0.639 |
| SEM (ML Error) | Queen contiguity | -533 | <0.001 | -645 | 0.025 | $\lambda = 0.487$ | <0.001 | 0.583 |

Notes: Values from CY 2022 analysis ( $N = 3,109$ counties). Outcome: county YPLL per 100,000 (CHR 2020-2022). All models control for social/economic factors, health behaviors, physical environment, and log population. Queen-contiguity $W$ constructed from Census county boundary file. SAR estimated via Maximum Likelihood (spreg ML\_Lag); SEM via ML\_Error. Neighbor exposure = proportion of $k = 5$ nearest counties with AI access.

### S10 Multi-Year Panel Comparison, 2022–2024

Table S19 summarizes cross-wave changes in technology diffusion, geospatial access, inequality, and county mortality model performance. The consistent negative AI coefficients across years and model types provide cross-wave replication. The divergence between coverage gains (+9.0 pp) and Gini deterioration (+0.028) indicates that new AI-enabled hospitals are disproportionately located in areas that already had relatively good access.

### S11 Covariate Balance Diagnostics and Overlap

We assessed the quality of the propensity score weighting by examining Standardized Mean Differences (SMD) before and after weighting. An SMD with an absolute value below 0.1 is often used as a rough heuristic, but in this setting the more important question is whether weighting compresses the largest confounders while the doubly robust outcome model still carries the lagged outcome and other residual imbalances directly.

**Table S19.**
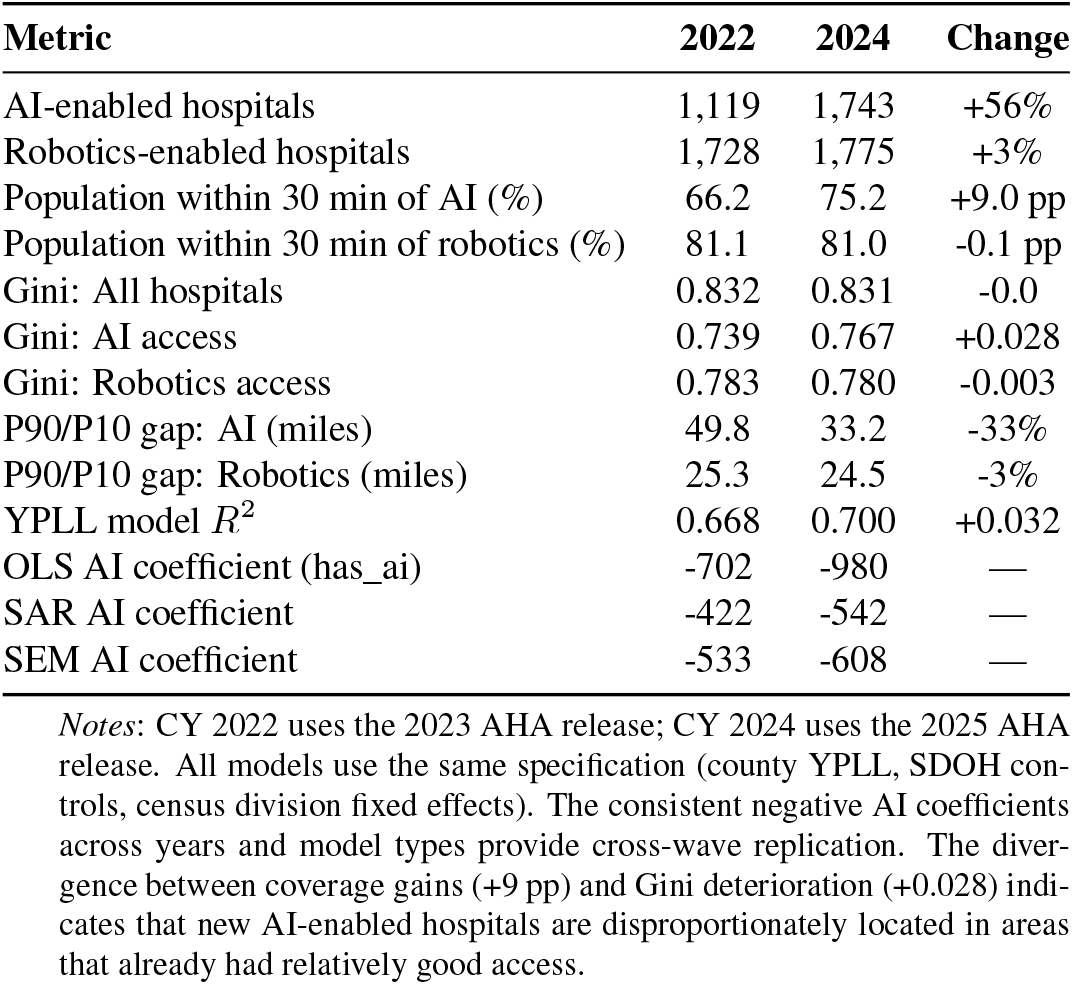
Multi-Year Comparison of Access and Inequality Metrics.

| Metric | 2022 | 2024 | Change |
| --- | --- | --- | --- |
| AI-enabled hospitals | 1,119 | 1,743 | +56% |
| Robotics-enabled hospitals | 1,728 | 1,775 | +3% |
| Population within 30 min of AI (%) | 66.2 | 75.2 | +9.0 pp |
| Population within 30 min of robotics (%) | 81.1 | 81.0 | -0.1 pp |
| Gini: All hospitals | 0.832 | 0.831 | -0.0 |
| Gini: AI access | 0.739 | 0.767 | +0.028 |
| Gini: Robotics access | 0.783 | 0.780 | -0.003 |
| P90/P10 gap: AI (miles) | 49.8 | 33.2 | -33% |
| P90/P10 gap: Robotics (miles) | 25.3 | 24.5 | -3% |
| YPLL model $R^2$ | 0.668 | 0.700 | +0.032 |
| OLS AI coefficient (has_ai) | -702 | -980 | — |
| SAR AI coefficient | -422 | -542 | — |
| SEM AI coefficient | -533 | -608 | — |

*Notes:* CY 2022 uses the 2023 AHA release; CY 2024 uses the 2025 AHA release. All models use the same specification (county YPLL, SDOH controls, census division fixed effects). The consistent negative AI coefficients across years and model types provide cross-wave replication. The divergence between coverage gains (+9 pp) and Gini deterioration (+0.028) indicates that new AI-enabled hospitals are disproportionately located in areas that already had relatively good access.

Fig. S2 illustrates the balance for the primary treatment variable: *AI for Automating Routine Tasks*. Prior to weighting (red circles), significant imbalances existed, particularly for population size, baseline YPLL, and neighboring adoption lags. After weighting (blue squares), the largest imbalances compress sharply toward zero, although the lagged outcome and health-behavior index remain modestly imbalanced—consistent with the main-text argument for retaining those terms directly in the outcome regression.

**Table S20.** Covariate balance: standardized mean differences before and after AIPW weighting (MO14 → DV21)

| Covariate | Unweighted SMD | Weighted SMD |
| --- | --- | --- |
| 2019 YPLL Rate | −0.630 | −0.222 |
| Log Population | 1.337 | 0.122 |
| Social/Economic Factors | −0.461 | −0.084 |
| Physical Environment | 0.206 | −0.158 |
| Health Behaviors | 0.111 | −0.248 |
| Medicaid Expansion | 0.347 | 0.090 |
| MO14 Spatial Lag | 0.726 | 0.099 |
| MO21 Spatial Lag | 0.527 | 0.082 |

*Notes:* SMD = standardized mean difference (treated minus control, divided by pooled SD). Census-division indicators and the remaining technology spatial lags are omitted for brevity; all were materially compressed after weighting. Residual imbalance on baseline YPLL and health behaviors motivates direct outcome adjustment in the doubly robust specification.

Table S21 provides diagnostics on the propensity-score weights and effective sample sizes (ESS) for the primary county estimand. Overlap was sufficient after clipping extreme weights at the 1st and 99th percentiles, and the overlap-weighted ATO estimates in Table S17 further reduce sensitivity to limited common support.

#### S11.1 SUTVA Diagnostics (Spillover and Interference)

To test the Stable Unit Treatment Value Assumption (SUTVA), we examined two potential violations: spatial spillovers (interference) and treatment variation.

- **Spillover Effects:** We compared outcomes for treated counties surrounded by high-adoption neighbors versus those surrounded by low-adoption neighbors. High-High (HH) clusters showed better raw outcomes (approx. 1,188 YPLL lower) due to baseline socioeconomic advantages, but High-Low (HL) “Islands” showed the strongest *adjusted* marginal association (*β* ≈ − 395), implying that spillovers likely contaminate the control group in clustered areas (conservative bias).
- **Treatment Heterogeneity:** We tested if the treatment effect varied by hospital size (Small vs. Medium vs. Large AI-enabled hospitals). ANOVA results indicated no significant heterogeneity (*p* ≈ 0.60), supporting our use of a binary treatment definition rather than an intensity-based metric, which was found to be collinear with urbanicity.

**Table S21.** Propensity-score and weight diagnostics for the primary MO14 → YPLL AIPW model.

| Quantity | Value |
| --- | --- |
| Counties ($N$) | 3,079 |
| Treated ($n$) | 416 |
| Control ($n$) | 2,663 |
| Treated prevalence | 0.135 |
| Propensity AUC | 0.871 |
| Propensity Brier score | 0.082 |
| Truncation rule | 1st / 99th percentile |
| Mean weight | 1.80 |
| Median weight | 1.07 |
| Weight 95th percentile | 4.49 |
| Weight 99th percentile | 14.23 |
| Max weight (before clipping) | 100.0 |
| ESS (treated) | 119 |
| ESS (control) | 2,376 |

Notes: ESS = effective sample size, calculated as $(\sum w_i)^2 / \sum w_i^2$ . Diagnostics are reported for the non-cross-fitted county estimand (MO14 → DV21). Cross-fitted ESS (409.1) and 99th-percentile weight (19.6) are reported in the main text (Balance and overlap diagnostics subsection).

### S12 Missingness and Reproducibility

Missing data was handled using listwise deletion for the primary analysis (*N* = 3, 143 counties with complete data). A sensitivity analysis using missingness indicators for key covariates (e.g., granular housing data) did not alter the direction or significance of the primary results. For hospital-level analysis, hospitals without valid 2024 CMS outcome data were excluded, but weights were recalculated to preserve representativeness. All code scripts and aggregated data files are provided in the GitHub repository to facilitate reproduction.

**Fig. S2.**
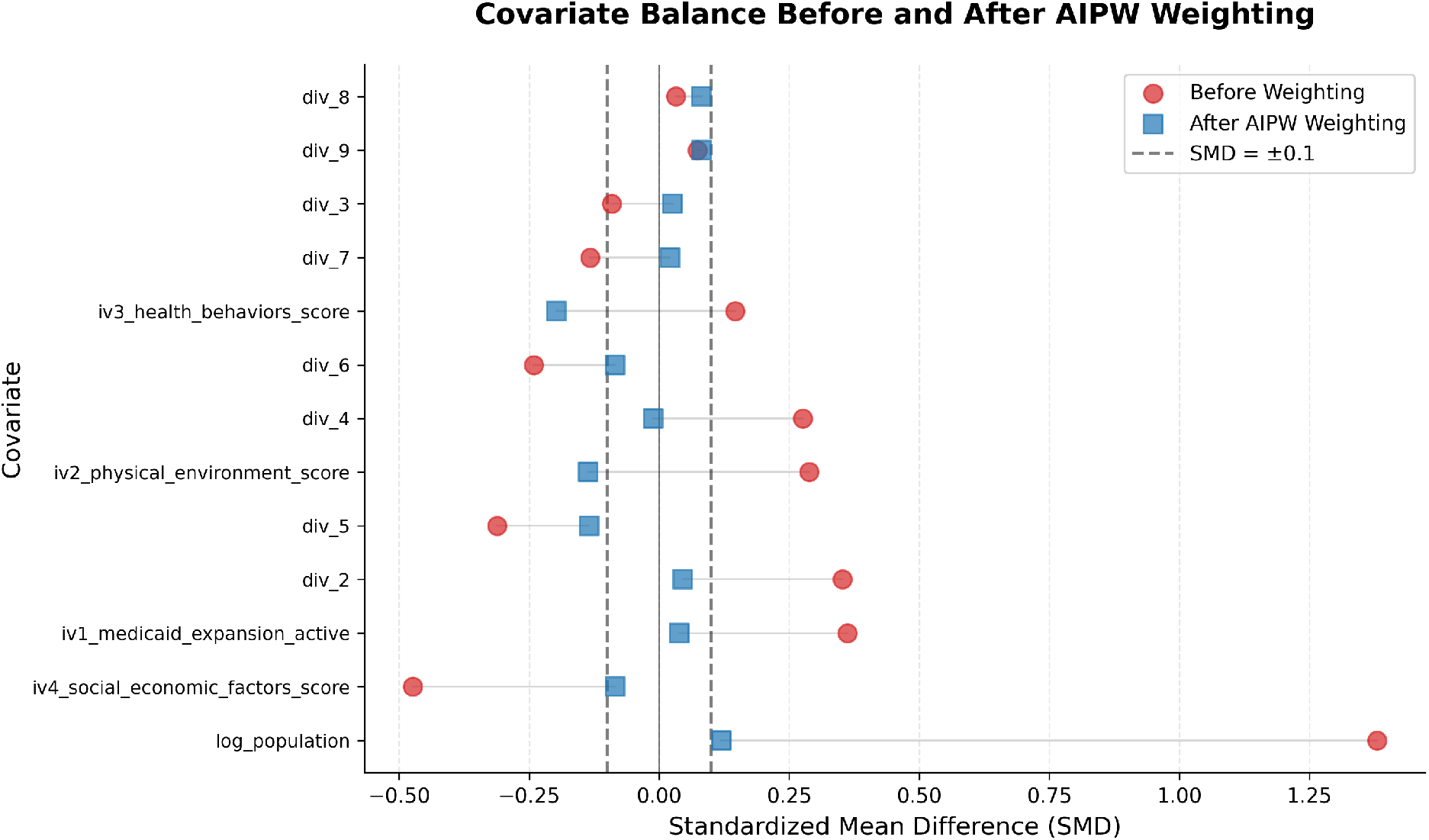
Covariate Balance (Love Plot) for AI Adoption. The plot displays the Standardized Mean Difference (SMD) between MO14-exposed and unexposed counties for key covariates. The red circles indicate the initial imbalance (unweighted), showing profound differences in population size, baseline mortality, and neighboring adoption. The blue squares indicate the balance after weighting. Most covariates move sharply toward zero, though baseline YPLL and health behaviors remain modestly imbalanced—one reason the doubly robust outcome model retains those terms directly. **Overlap diagnostics:** companion diagnostics in Tables S20–S21 summarize balance, effective sample sizes, and weight tails.

